# Evidence Supports a Causal Role for Vitamin D Status in Global COVID-19 Outcomes

**DOI:** 10.1101/2020.05.01.20087965

**Authors:** Gareth Davies, Attila R Garami, Joanna Byers

**Affiliations:** Independent Researcher

## Abstract

**Background:** The COVID-19 pandemic caused by the coronavirus SARS-CoV-2 seemed to affect locations in the northern hemisphere most severely appearing to overlap with the pattern of seasonal vitamin D deficiency. Integrating available knowledge, we hypothesised that vitamin D status could play a causal role in COVID-19 outcomes.

**Objectives:** We set out to analyse the relationship between COVID-19 severity and latitude, and construct a causal inference framework to validate this hypothesis.

**Methods:** We analysed global daily reports of fatalities and recoveries from 239 locations from 22nd Jan 2020 to 9th April 2020. We quantified local COVID-19 outbreak severity to clearly distinguish the latitude relationship and identify any outliers breaking this pattern, and analysed the timeline of spread. We then used a causal inference framework to distinguish correlation from cause using observational data with a hypothetico-deductive method of proof. We constructed two contrasting directed acyclic graph (DAG) models, one causal and one acausal with respect to vitamin D and COVID-19 severity, allowing us to make 19 verifiable and falsifiable predictions for each.

**Results:** Our analysis confirmed a striking correlation between COVID-19 severity and latitude, and ruled out the temporal spread of infection as an explanation. We compared observed severity for 239 locations with our contrasting model. In the causal model, 16 predictions matched observed data and 3 predictions were untestable; in the acausal model, 14 predictions strongly contradicted observed data, 2 appeared to contradict data, and 3 were untestable.

**Discussion:** We show in advance of RCTs that observed data strongly match predictions made by the causal model but contradict those of the acausal model. We present historic evidence that vitamin D supplementation prevented past respiratory virus pandemics. We discuss how molecular mechanisms of vitamin D action can prevent respiratory viral infections and protect against ARDS. We highlight vitamin D’s direct effect on the renin-angiotensin-system (RAS), which in concert with additional effects, can modify host responses thus preventing a cytokine storm and SARS-CoV-2-induced pathological changes. Emerging clinical research confirms striking correlations between hypovitaminosis D and COVID-19 severity, in full alignment with our study.

**Conclusions:** Our novel causal inference analysis of global data verifies that vitamin D status plays a key role in COVID-19 outcomes. The data set size, supporting historical, biomolecular, and emerging clinical research evidence altogether suggest that a very high level of confidence is justified. Vitamin D prophylaxis potentially offers a widely available, low-risk, highly-scalable, and cost-effective pandemic management strategy including the mitigation of local outbreaks and a second wave. Timely implementation of vitamin D supplementation programmes worldwide is critical with initial priority given to those who are at the highest risk, including the elderly, immobile, homebound, BAME and healthcare professionals. Population-wide vitamin D sufficiency could also prevent seasonal respiratory epidemics, decrease our dependence on pharmaceutical solutions, reduce hospitalisations, and thus greatly lower healthcare costs while significantly increasing quality of life.

## Introduction

In the ongoing COVID-19 pandemic, significant numbers of fatalities appeared to occur in the northern hemisphere with far fewer in the tropics and southern hemisphere. Past coronavirus and influenza epidemics and pandemics have displayed very strong seasonality [1]. Seasons are determined by latitude, and winter provides a more favourable environment for viral transmission: enveloped viruses are fragile and more easily destroyed by heat and UV light than other viruses [2]. Therefore, one would expect transmission rates to vary by latitude, however, overall COVID-19 data suggest that case fatality rates (CFRs) also appeared to be strongly governed by latitude, and this is highly unexpected.

A possible causal role for vitamin D in global COVID-19 outcomes was proposed as early as 12th March 2020 [3] by Dr Garami, and the idea began to be discussed and gain support [4]. The authors had simultaneously been researching this area and joined to publish a summary of early findings for medical professionals on 19th March [5], calling for hospitals to begin to treat, test, measure and report. Vitamin D is a logical candidate hypothesis since it is produced in the skin by sufficient sunlight and its deficiency is latitude- and season-dependent [6][7]. Vitamin D is a steroid hormone with receptors in most tissue cell types [8], regulating multiple biological pathways, and vitamin D deficiency is associated with increased risk for chronic diseases including autoimmune diseases, many cancers, cardiovascular disease, infectious diseases, dementia, schizophrenia and type 2 diabetes [8,9]. Furthermore, Italy, one of the worst countries affected by COVID-19 in Europe from February through April, has a high prevalence of vitamin D deficiency [10].

Known risk factors for low vitamin D status are old age, winter, living at higher latitudes, darker skin pigmentation, less sunlight exposure, dietary habits, obesity, malabsorption syndromes and absence of vitamin D fortification [11][12].

COVID-19 fatalities in BAME communities have been unproportionally high in the UK with over a third of ICU admitted [13,14] and 63.2% (67/106) of the first 106 health and social care professionals had BAME background [15]. The New York Department of Health and Mental Hygiene also reported that Black/African American and Hispanic/Latino populations are suffering up to twice the case fatality rates of white New Yorkers [16].

Mounting evidence indicates a correlation between vitamin D and all-cause morbidity, all-cause mortality, respiratory infections, acute respiratory distress syndrome (ARDS), as well as with comorbidities associated with COVID-19 such as diabetes mellitus [17]. Some studies have inferred a causal role with MS and colorectal cancer [18]. Contradicting results and methodology concerns raised the question of whether vitamin D plays a causal role or is simply a bystander, and is merely a biomarker for overall disease-health status.

Given the difficulties of running large randomized controlled trials (RCTs), more recent efforts have turned to Causal Inference methods. Modern techniques such as Mendelian Randomisation allows the question of causality to be investigated using observational data sets. A meta-analysis of European cohort studies used Mendelian Randomisation to provide further support for a causal relationship between vitamin D deficiency and increased all-cause mortality. However, the study size, (~10,000 participants), was considered underpowered and larger studies on genetics and mortality were called for [19]. Meta-analyses showing causal relationships between vitamin D supplementation and morbidity and mortality exist. Intake of ordinary doses of vitamin D supplements seems to be associated with decreases in total mortality rates [20]. An RCT of higher doses of vitamin D supplementation showed no reduction in total cancer incidence, however vitamin D significantly reduced the total cancer mortality [21].

The worldwide scale of the COVID-19 pandemic provides a unique opportunity to apply high-level causal inference models with a wide set of independent root causes (“exogenous variables”). This enables a simple but powerful *qualitative* analysis of model predictions allowing causal inference by comparing predictions with observed data. Randomisation in Causal Inference is performed by nature rather than the experimenter, but non-randomised observational data can also be used to infer causality using Bayesian analysis to remove selection bias [22].

Our study objectives are two-fold. First, we analyse and quantify severity of global COVID-19 outbreaks to define trends and outliers. Secondly, we apply a causal inference framework to answer the binary question: has vitamin D played a causal role in outbreak severity?

Readers unfamiliar with Causal Inference may find it useful to first read *Appendix 1 - Background on Causal Inference*.

## Methods

### Data Sources, Processing and Code

COVID-19 data on cases, recoveries and deaths was provided by John Hopkins CSSE’s public github repository [23]. Some CSV files were converted to Excel format for previsualisation checks before being imported into Matlab. Inconsistencies in location definitions between deaths and recoveries data for Canada^1^ were reconciled before processing and analysis of the data.

Data for population by latitude was provided by the Center for International Earth Science Information Network, Columbia University [24].

All code, data and figures used in this analysis are available online in a github repository [25].

### Methods 1 - Analysis Of Severity Versus Latitude and Time

We analysed COVID-19 fatalities by latitude up to 28th March 2020 aggregating total deaths for each location into data bins of size five degrees each over the range 40°S to 70°N. We aggregated world population latitude data into corresponding bins and calculated case fatalities per million as a function of latitude.

We compared severity by location with the known timeline of spread of COVID-19 around the globe to show that timing of infection was not involved in determining outbreak severity.

We then performed a detailed analysis of outbreak severity by location for all 239 global reporting locations over the same period. Outbreak severity was judged using the ratio of reported recoveries to deaths and the total number of deaths. We calculated the Epidemic Severity Index (ESI) [26] with a reference severity defined by a hospital survival ratio, *S*, of 6.5, corresponding to a disease severity approximately twice that of season flu in the US.

We plotted ESI scores using geobubble plots using both linearised and log scales. In the linearised ESI plots bubble area is directly comparable to the total number of deaths. The logarithmic ESI form is used to show locations with severity index values too small to be seen on the linearised plots.

We considered all locations that reported infections within seven weeks of the initial outbreak in Hubei to identify outliers breaking the latitude dependence pattern for detailed analysis. We manually included some locations infected outside seven weeks that appeared worthy of more detailed analysis.

### Methods 2 - Causal Inference Model Analysis

In order to explain the results of the latitude analysis in section one and additional available global data and literature relating to COVID-19 outcomes we use a formal causal inference (CI) framework and model causal relationships using directed acyclic graphs (DAGs).

In medicine, causal models are normally the basis for inverse analyses^2^, starting with the data and using model-based statistics to estimate average treatment effect sizes. Such methods require large, reliable data sets of variables. No such data set was available ruling out an inverse analysis. However, the vast data set that was available was highly suited to a forward method. Forward methods start with the model and generate testable predictions about the data we would expect to observe. This is a classic example of the hypothetico-deductive method, one of the earliest, most powerful and successful scientific methods.

To create our DAG models, we first categorised variables by their expected causation effect to simplify the model relationships. We listed specific members of each category in order to highlight possible confounders that would otherwise potentially be masked by this categorisation.

Inverse methods construct a single model, but in our forward method analysis we sought the answer to a binary question - whether vitamin D plays a causal role in COVID-19 disease outcomes or not - and so we expressed the causal/not-causal dichotomy with two contrasting models (Figure 1 and Figure 2). Technically, these are two views of the same model with binary values for the strength of the causal connection. The two models generated different testable predictions about the outcomes we should expect if each were true.

**Figure 1.**
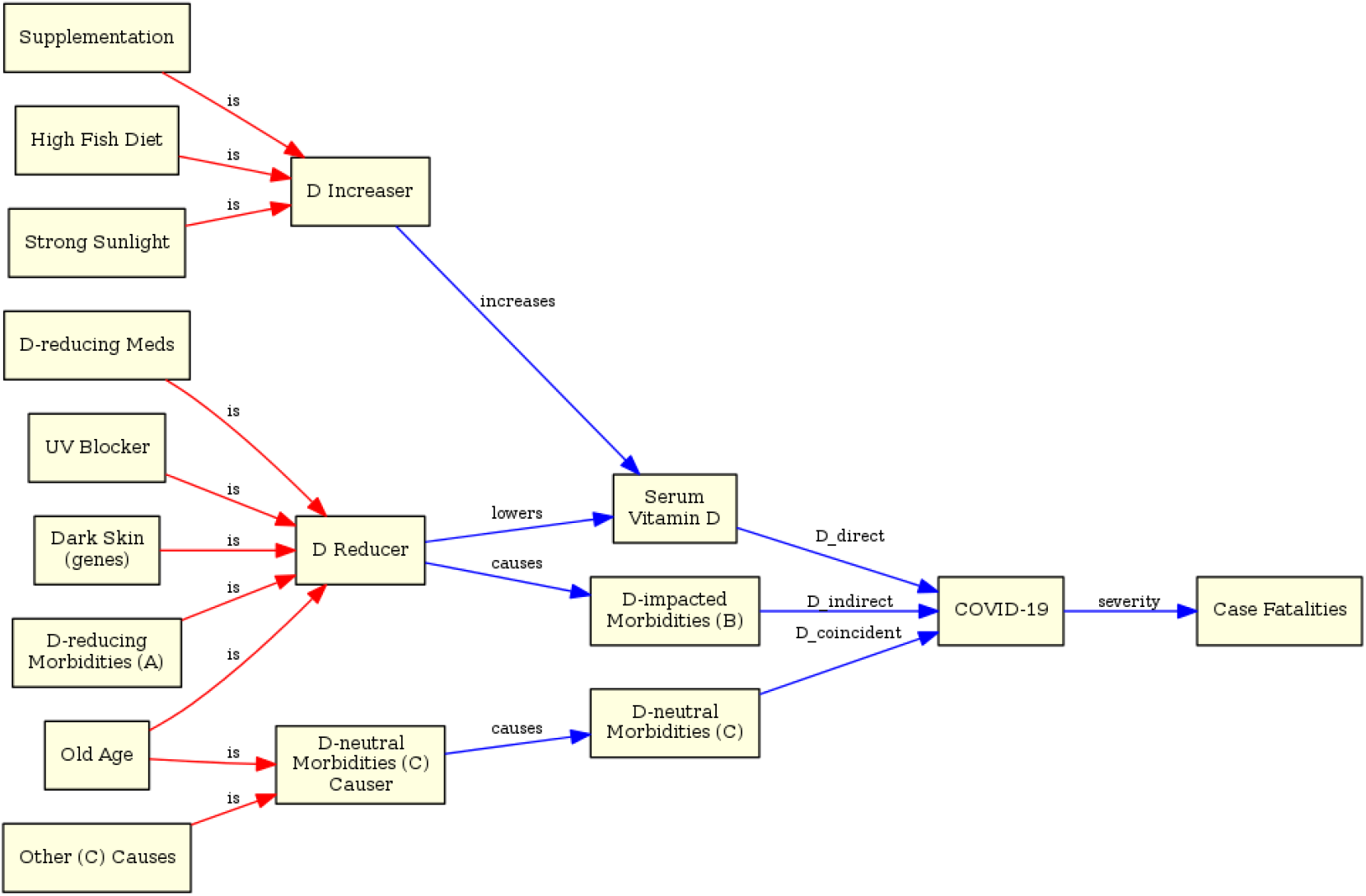
Model A (casual) - serum Vitamin D levels play direct and indirect (“mediated”) causal roles in disease severity and outcomes.

**Figure 2.**
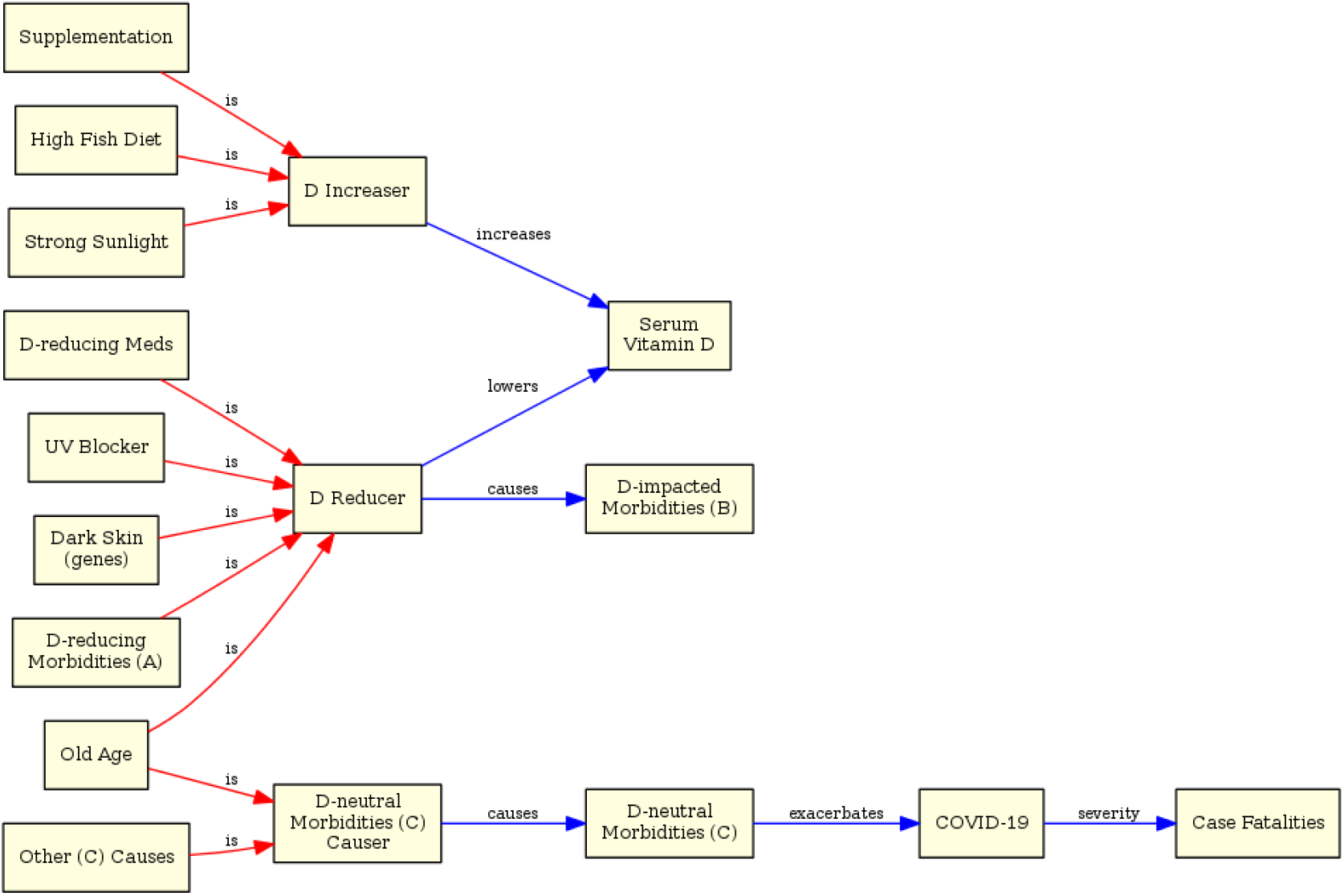
Model B (acausal or “bystander”) - direct and mediated causal links between serum Vitamin D levels and COVID-19 have been removed.

We used three categories for exogenous variables (root causes) influencing COVID-19 outcomes:

1. Sources of vitamin D (“D Increasers”)
2. Causes of vitamin D deficiency (“D Reducers”)
3. Causes of Morbidities with no vitamin D relationship (“D-neutral Morbidities Causes”)

We categorised morbidities in a similar positive/negative/neutral sense:

A. “D-reducing” (illnesses that lower vitamin D serum levels)
B. “D-impacted” (illness caused by or exacerbated by low vitamin D levels)
C. “D-neutral” (no causal relationship with serum vitamin D levels)

We included a (non-exhaustive) set of examples for each variable category with lines drawn in **red**^3^ to keep them visually distinct from the causal paths of interest in **blue**. Most of these variables are self-evidently independent.

Note that since “D-reducing Morbidities” is an exogenous variable, it appears in the DAG model in the set of examples for the “D Reducers” category.

We analysed the model for potential biases and confounders and then evaluated the predictions made by both models, tabulating these to highlight differences.

Finally, we cross-referenced available observed data and compared these with model predictions.

## Results

### Results 1 - Analysis Of Severity Versus Latitude and Time

#### Severity by Latitude

COVID-19 fatalities display a striking correlation with latitude. Figure 3 shows fatalities by latitude, human population by latitude, and finally fatalities per million people by latitude. Severe outbreaks with large fatalities occurred exclusively above the +30°N latitude line, in the winter hemisphere, where fatalities per million ranged from 3% to 37% (mean 11%) for latitudes in the range 30°N to 55°N. Outbreaks in the tropics and southern summer hemisphere were very mild with an average of just 0.2% fatalities per million.

**Figure 3.**
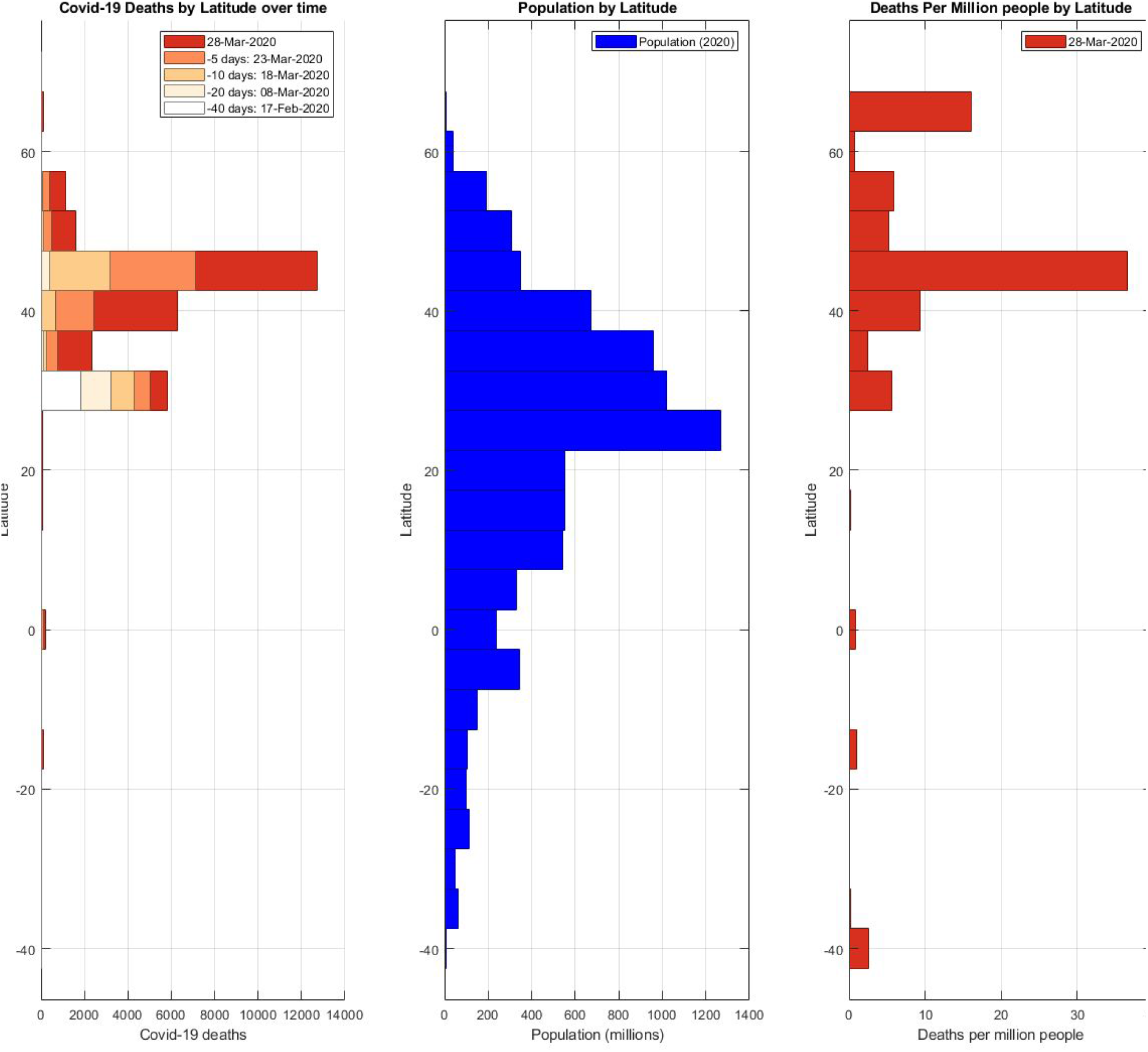
(left) COVID-19 fatalities by latitude and over time; (middle) 2020 population by latitude; (right) COVID-19 fatalities per million people by latitude. Note: the Deaths per Million value at -40°S is a statistical artefact due to dividing two small numbers and may be ignored.

The timeline of spread of infection is shown in Figure 4.

**Figure 4.**
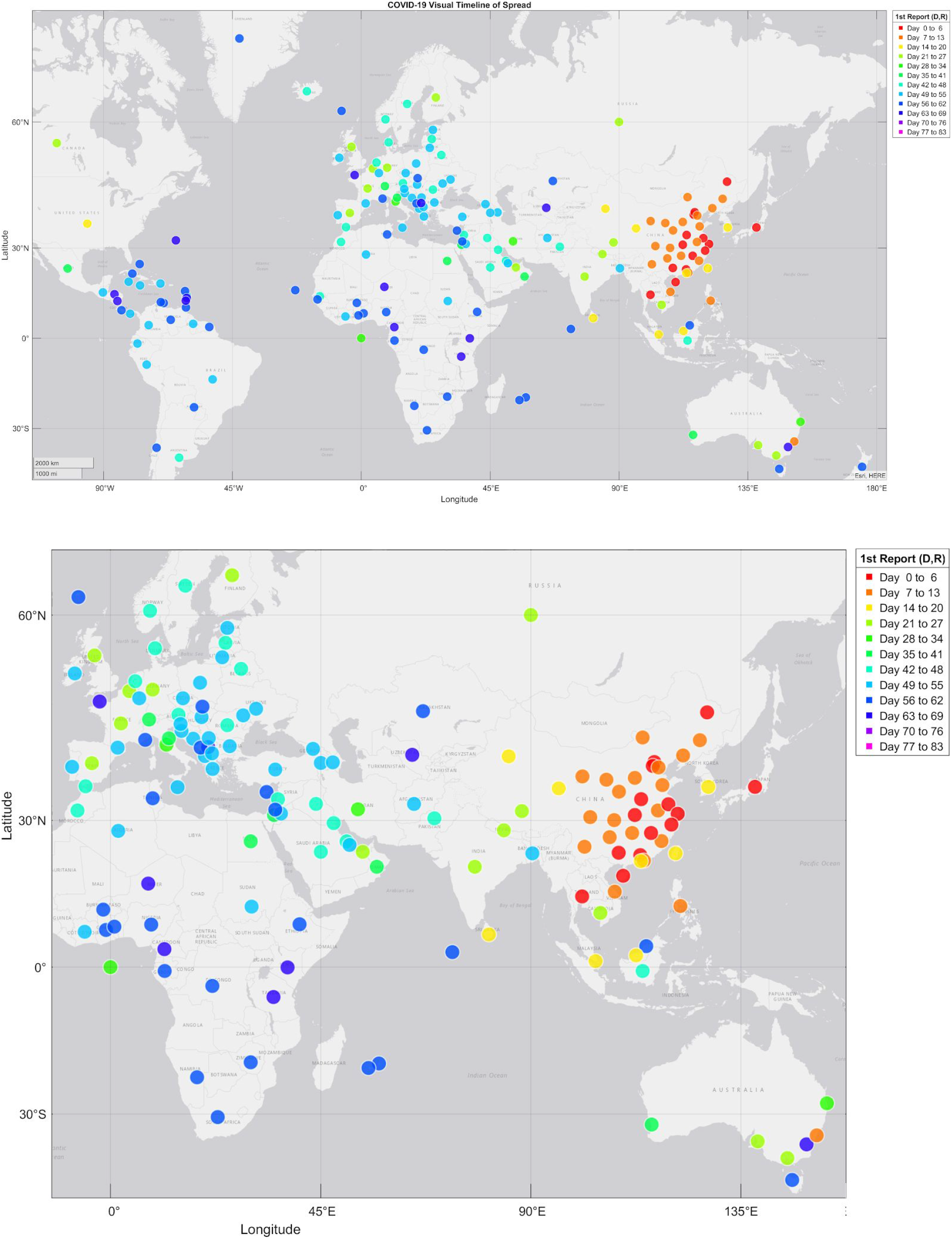
(top) Visual timeline of global spread of COVID-19 (first reported death or recovery), (bottom) magnified section showing Europe, Africa, Asia and Oceania.

#### Quantifying Local Epidemic Severity

In order to compare the timeline of spread with specific locations, we calculated the Epidemic Severity Index (ESI) for all 239 reporting locations. We conditioned the ESI to judge severity against a reference baseline survival ratio, *S = 6.5*, which can be interpreted as meaning any outbreak that is worse than approximately “twice as severe as US seasonal flu”.

Figure 5 shows peak ESI scores (linearised form^4^) for all locations up to the end of March 2020 marking the end of winter and the start of spring. Nine locations experienced severe outbreaks over the period (ESI > 2.5), all of them above the 30°N latitude line.

**Figure 5.**
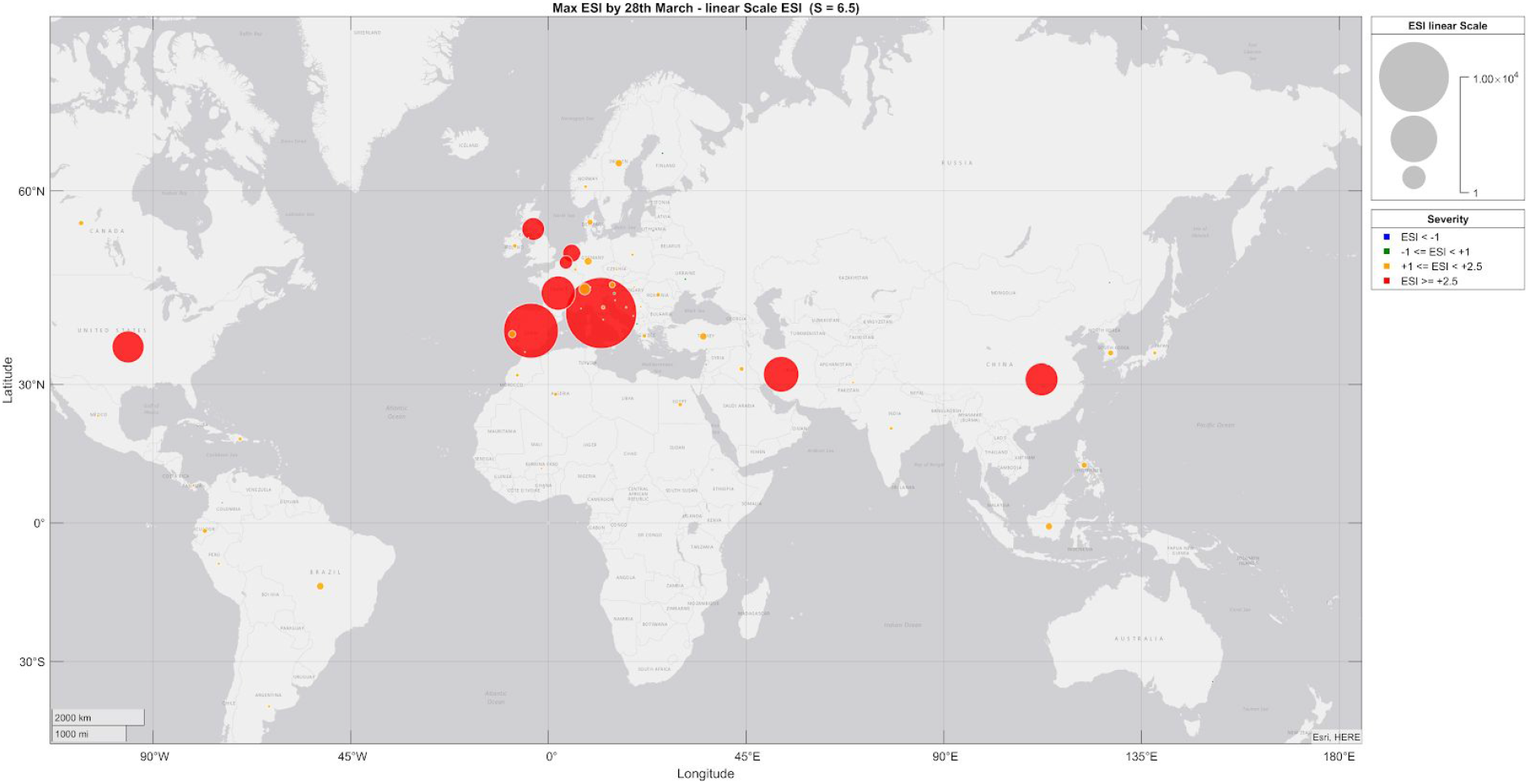
peak ESI scores up to 28th March 2020 (S = 6.5; linearised scale) Colour Key: Yellow = ESI < +2.5 (mild); Red = ESI > +2.5 (severe)

Figure 6 shows the log scale version of the same data to show low severity outbreaks more clearly - note that some location’s ESI scores went negative immediately and therefore do not show since their peak ESI is 0.

**Figure 6.**
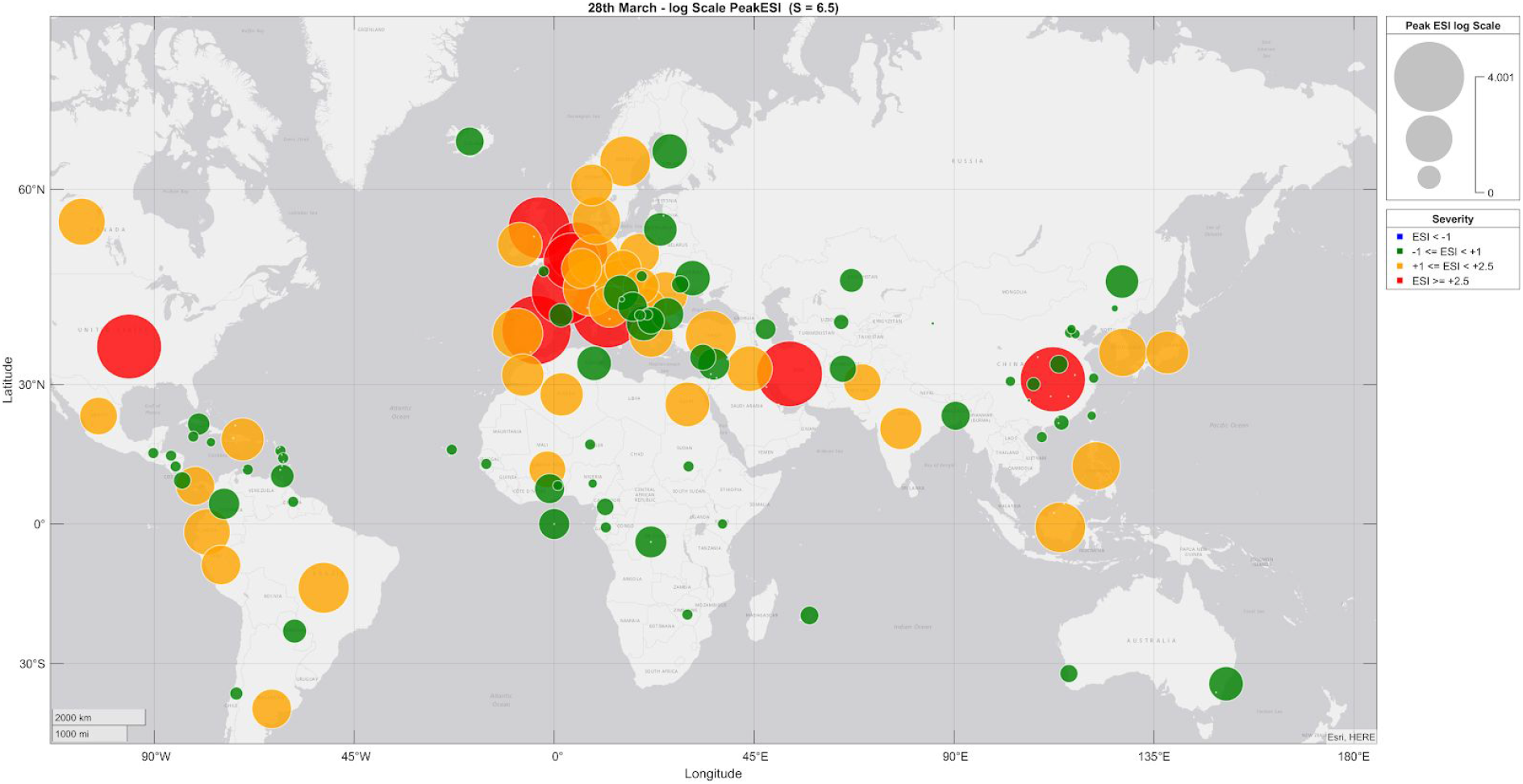
peak ESI scores for all locations up to 28th March 2020 (log scale) Colour Key: Green = ESI < +1 (very mild); Yellow = +1 < ESI < +2.5 (mild / not yet severe); Red = ESI > +2.5 (severe)

Table 1 is a shortlist of 35 locations for detailed analysis. The general criteria for inclusion in the shortlist was, other than Hubei, locations outside China that showed evidence of infection within the first 7 weeks (+48 days) after reporting began (22nd January). We selected four more locations outside this cutoff date where ESI values indicated they might be outliers or of particular interest for analysis. Peak and final ESI scores are shown along with details of total deaths and recoveries by 28th March (+66 days). We calculate an approximate value for hospital fatality rate^5^ for locations where there was sufficient data (N>99) for this to be minimally significant. This approximation assumes that early recoveries and deaths were reported by hospitals. This is reasonable since tests were not widely available during this early period therefore hospital deaths and discharges would be the predominant source of early reported figures and this assumption is sufficiently robust to make meaningful comparisons of relative severity for early infections. Data for Canada required special reconciliation and is marked grey.

**Table 1.**
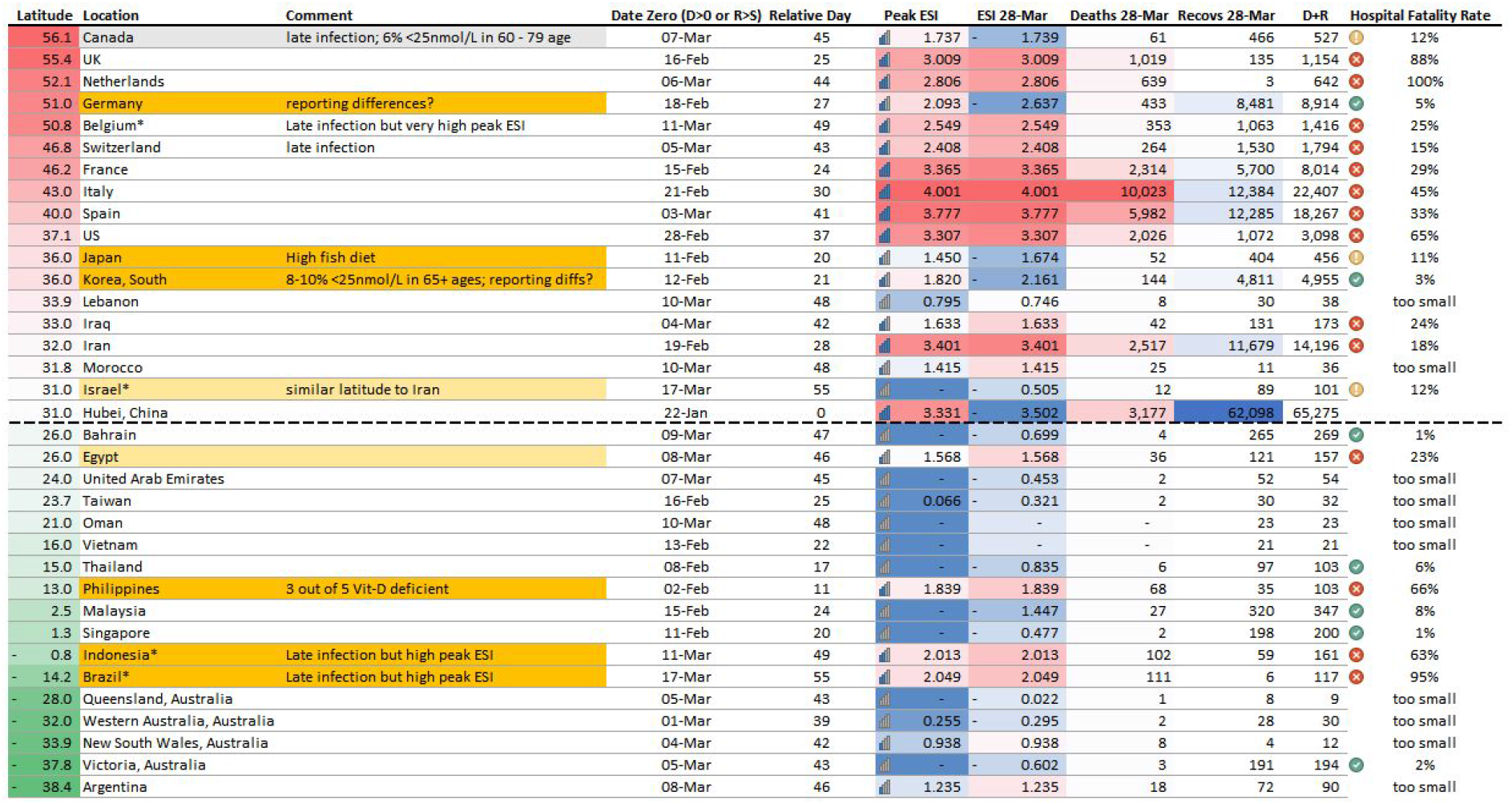
Shortlisted locations infected within 48 days, plus manually selected locations of interest (marked *). Pattern outliers marked in orange. Peak ESI and latest ESI up to 28th March show severity. Death, Recoveries, Totals and approximate Hospital Fatality Rates.

We judged the date of first infection (“day zero”) for each location using a heuristic criterion consistent with our ESI calculations: that is, either the date of the first reported death or the date where reported recoveries exceeded *S*, whichever condition was met first (*S* = *6.5*).

Outliers breaking the latitude dependency are marked in orange. Light orange indicates possible outliers. Hand-selected locations of interest are marked with an asterisk.

The latitude dependency of outbreaks is shown by Figure 7 and Figure 8 which plot the Epidemic Severity Index over time for all locations shortlisted in Table 1. Locations were grouped in descending order of latitude so that each subplot contained a maximum of six locations for readability.

**Figure 7.**
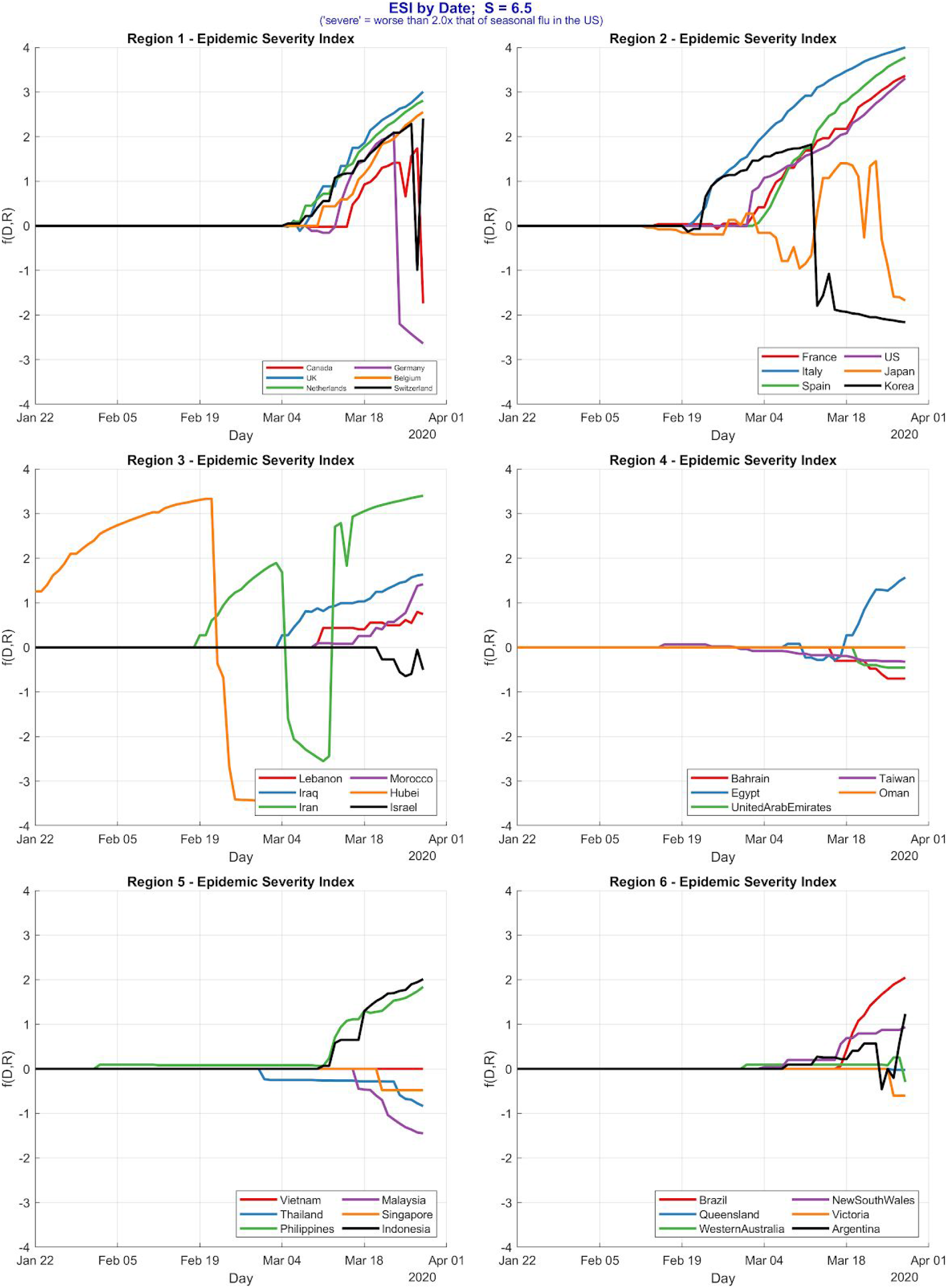
Epidemic Severity Index (*S* = *6.5*) by date for all locations infected within 7 weeks of Hubei, plus locations grouped into six regions by descending order of latitude. (ESI > 2.5 is severe).

**Figure 8.**
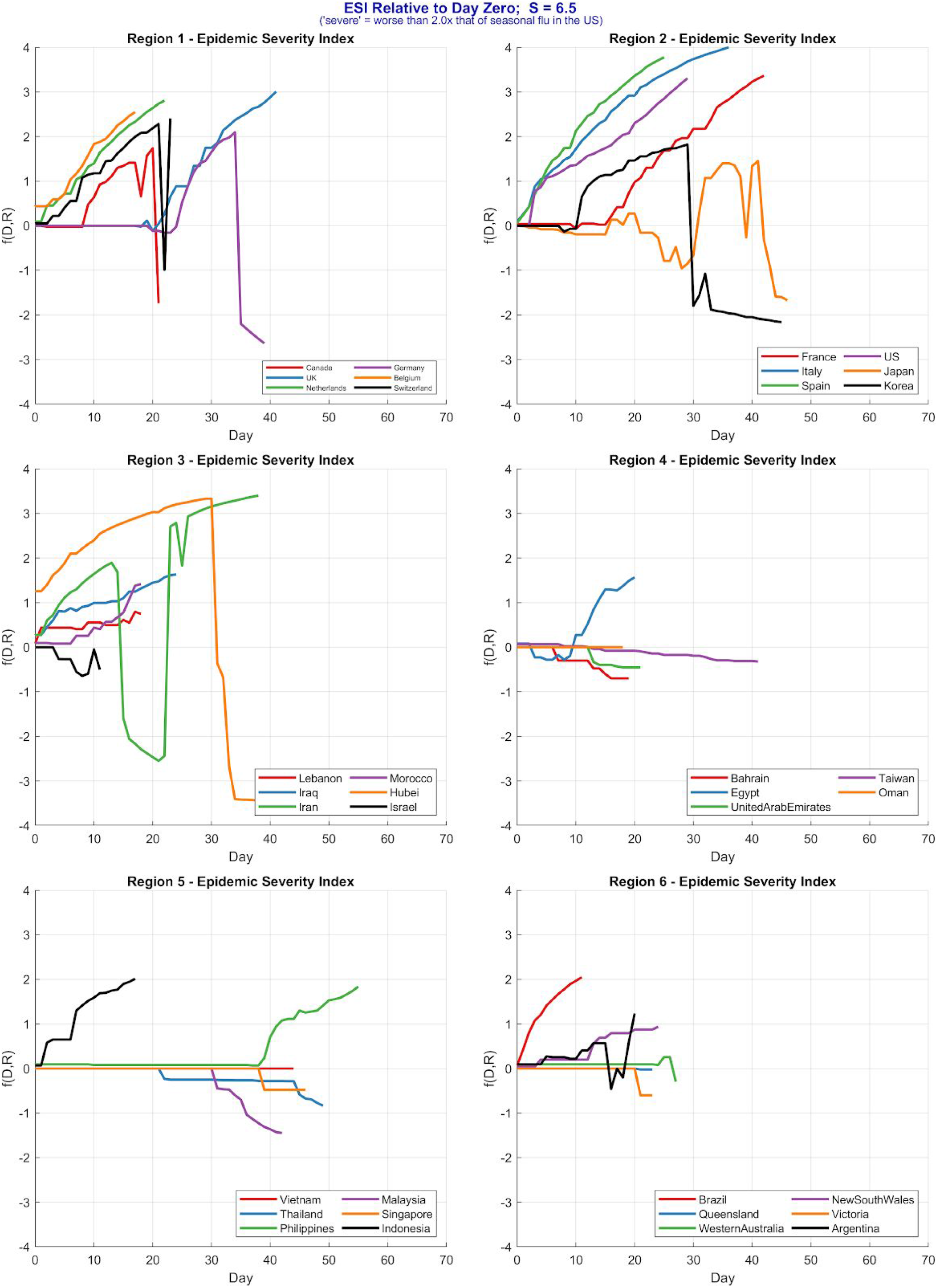
ESI (*S* = *6.5*) relative to “day zero” for locations reporting within 7 weeks of Hubei, plus selected locations, grouped into six regions in descending order of latitude. (ESI > 2.5 is severe).

Figure 7 shows ESI by actual date. Figure 8 shows ESI by relative day number counted from each location’s “day zero”. It should be noted that ESI and timeline of spread assume that fatalities and recoveries figures relate to local populations and not travellers. This assumption is not always valid and for some locations the “day zero” may appear shifted earlier in time than the true day zero since travellers were infected in a different location. This shift is most visible in severe outbreaks where the outbreak begins visibly to rise some weeks after day zero when the true local infection takes hold. We have not adjusted for this since it is clearly visible in the timeline ESI plots in each case and we did not require a more rigorous definition of “day zero” for this analysis.

Although not identified in Table 1, we included Israel in the ESI timeline plots and analysis since it reported infections within 42 days but had very low deaths despite having latitude comparable with Iran which suffered a more severe outbreak typical of locations above 30°N.

#### Identification Of Outliers

Five locations above 35°N appear as pattern outliers with relatively mild outbreak severities: Canada, Switzerland, Germany, Japan and South Korea.

Iran shares a latitude comparable with Israel and while Israel did not match our crude initial criteria for determining outliers, a more detailed look at the data suggested comparing the two locations would be worthwhile.

In the region below 30°N, the Philippines appears to be an obvious outlier and possibly Egypt though its later infection makes this less clear. Other locations in the south that were infected later but show trends suggestive of more severe outbreaks include Brazil and India.

### Results 2 - Causal Inference Model Analysis

#### Confounder Bias and Special Cases

In general, any “upstream” causes of forks may be sources of confounding in observational data. Given the broad nature of causes of morbidities, this can cause complexity (Figure 9). This can be handled by considering specific morbidities and controlling for common causes, or employing alternative decounding mechanisms. Modern techniques, such as the “front door criterion” enabled deconfounding of even unknown or unmeasured variables through the use of mediator variables.

**Figure 9.**
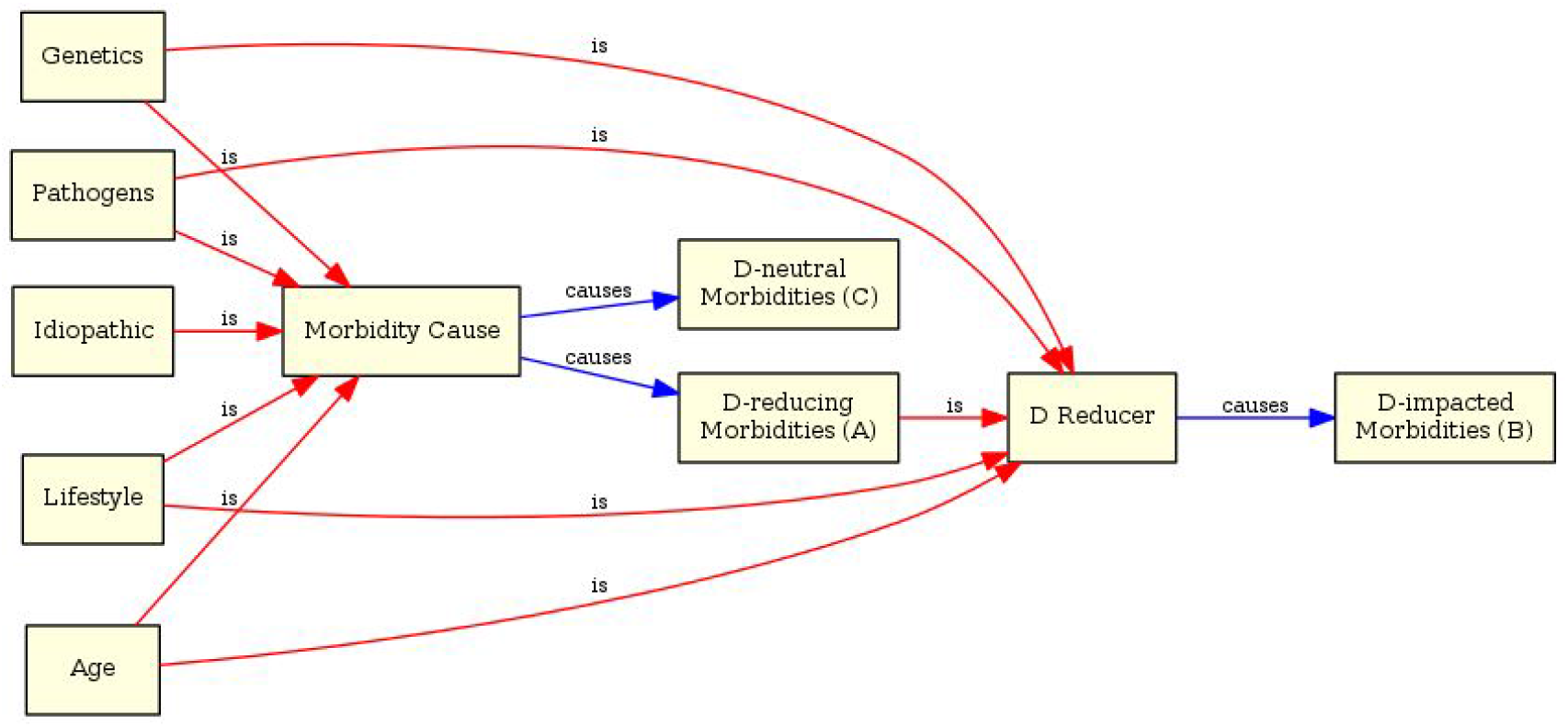
upstream causes must be controlled for when analysing morbidity statistics

Another way to reduce bias is triangulation. Triangulation integrates results from different data where each set has different sources of potential bias that are unrelated. If these different data all point to the same conclusion, this strengthens confidence in the finding [27].

#### Special Cases: Senescence, Pregnancy and Infancy

In developed countries, pregnant women and infants are routinely supplemented with vitamin D [28], so in Model A we consider these as belonging to the Supplementation “D increaser” root variable category.

In model B, where vitamin D is not causal, we consider them as part of “D-neutral Morbidities”, expanding the condition of “morbidity” to include the immunosuppressed state of pregnancy and still-developing states of infant immune systems.

Other modelled root causes that we shall consider are, to all intents and purposes, independent.^6^ I.e. we expect that taking supplements has zero effect on skin colour genes, and does not change the orientation of the Earth with respect to the sun *etc*. The special case of season impacting vitamin D supplementation is *not* confounding since the effects compensate for each other in terms of impact on outcome (which is the point of taking supplements during winter). However, since we will be considering specific cases of season in our qualitative analysis, this potential confounder will be “controlled for” anyway.

#### Cyclic Dependencies

Pathogen-caused disease spread has a cyclic nature: pathogens cause infections which cause disease which results in more pathogens (Figure 10). Cyclic dependencies can’t be handled by DAG causal models which are “acyclic” by definition. The acyclic restriction is necessary for quantitative analysis because cyclic dependencies (positive or negative feedback loops) are mathematically very complex to model^7^. However they can be analysed qualitatively and the relationship is worthy of discussion.

**Figure 10.**
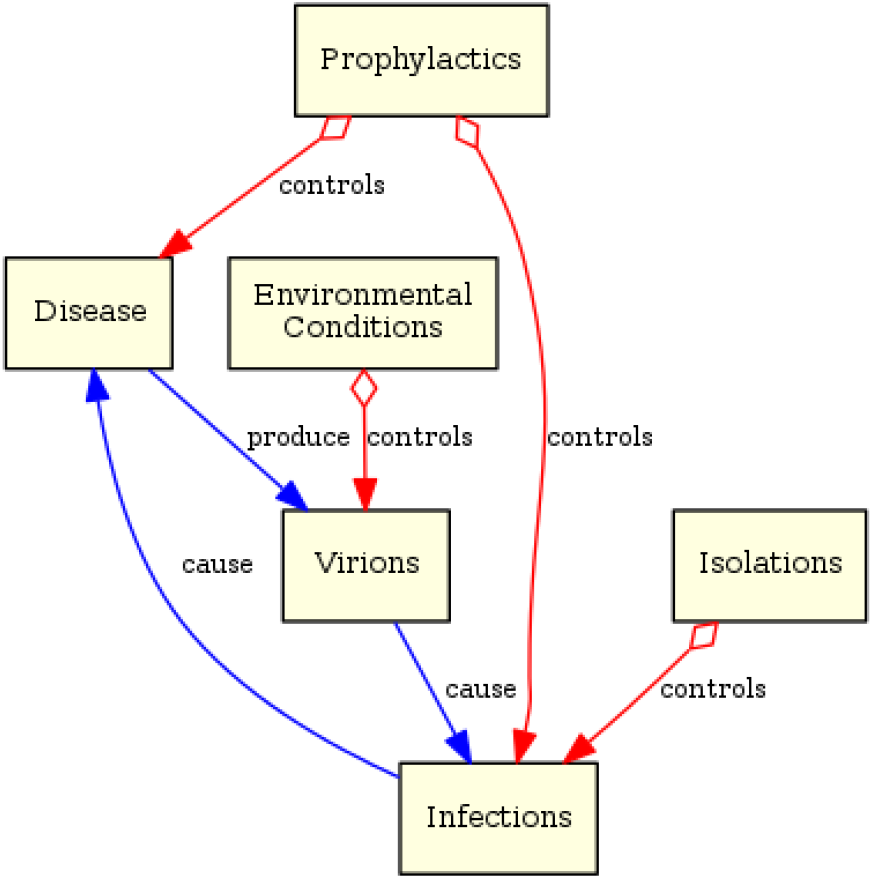
probability of spreading and catching viral diseases have cyclic dependencies

It’s clear from the viral cycle that anything that controls disease severity, or reduces the number of viral particles, or lowers the probability of infection, will suppress the feedback loop.

Environmental conditions hostile to virus particle survival will reduce transmission rates; Isolation, quarantine and social distancing measures act similarly but neither is expected to reduce disease severity or case fatalities rates (CFRs).

Anything that directly controls disease severity will act to reduce CFRs as well as reducing transmission rates since fewer viral particles are produced and this further reduces the possibility of infection. Prophylaxis will therefore lower infection rates as well as CFRs. This is an important distinction.

Informed by these considerations and armed with two contrasting models we can now make predictions for each.

### Causal Model Predictions

Model A and Model B predict multiple different outcomes. A small number of predictions made by the two models are the same and so these cannot be used to answer the question of causality.

Season is a function of latitude and can only take on one value in any given location at a given time. Since D reducers and D increasers act in opposition, we consider the cases of D increasers counteracting the D-reducing effects of weak sunlight (winter latitudes); and D reducers counteracting the D-increasing effects of strong sunlight (summer and tropical latitudes).

#### Causal Model A Predicts

1. Latitude will correlate strongly with CFRs, with winter locations showing worse CFRs than tropical and summer locations.
2. Winter locations with atypically low prevalence of vitamin D deficiency will appear as outliers with unusually low transmission and CFRs - even in the absence of quarantine & isolation measures.
3. In winter locations, typically high-risk subpopulations that are routinely supplemented with vitamin D will stand out as unexpectedly low-risk: pregnancy and infancy.
4. In winter locations, subpopulations with dark skin will correlate with higher CFRs.
5. Tropical and summer hemisphere locations will in general display lower transmission and CFRs - even in the absence of quarantine & isolation measures.
6. Tropical and summer hemisphere locations with atypically high prevalence of vitamin D deficiency - especially in elderly populations - will appear as outliers experiencing higher levels of transmission and case fatalities.
7. All D reducers will correlate with higher CFRs even in the absence of any obvious comorbidities.
8. D-reducing Morbidities (A) will correlate with CFRs.
9. D-impacted Morbidities (B) will correlate with CFRs even when controlled for confounder bias.
10. In tropical and summer locations, populations with high prevalence of D deficiency from e.g. extreme sun-blocking or avoidance behaviours - such as Islamic women wearing full body-covering clothing - will correlate with higher CFRs (note that facial clothing may act to lower transmission rates however metrics for transmission rates for subpopulations were not available to this study).

#### Bystander Model B Predicts

1. Latitude will not correlate with CFRs.
2. In winter locations, vitamin D sufficiency, and high-fish diets will not correlate with lower CFRs.
3. Routinely supplemented subpopulations will not correlate with CFRs: pregnancy and infancy will show up as typically high-risk.
4. D reducers will not correlate with CFRs in the absence of illness.
5. In winter locations, only quarantine, isolation and social distancing measures will correlate with lower transmission rates.
6. Only D-neutral Morbidities (C) will correlate with CFRs once disease confounders are controlled for.

### Predictions Compared to Available Data

We use total hospital cases per location (Deaths + Recoveries) (see Table 1) as a proxy indicator of transmission rates, and Hospital Fatality Rates (D/(R+D)) as a proxy indicator of CFR. Table 2 summarises the predictions from each model, grouping broadly by latitude to consider counterations and lastly a general predictions group, and compares predictions (P) with observations (O). Table cells have been coloured to clearly indicate where predictions match observations: green = strong match; orange = mismatch; red = strong mismatch. Predictions that are the same in both models are coloured grey as these are not informative. Cells where predictions cannot be validated against observations are left white.

**Table 2.** Predictions from models A and B. Cases where the models predict the same outcome have been coloured grey.

| Variables | Causal Model A |  | Bystander Model B |  |
| --- | --- | --- | --- | --- |
| Winter Latitudes > 40°N | Transmission | CFRs | Transmission | CFRs |
| General Winter Populations | P: higher<br>O: higher | P: higher<br>O: higher | P: higher<br>O: higher | P: baseline<br>O: higher |
| Quarantine, Distancing Measures | lower | P: same as general (higher)<br>O: higher | lower | P: same as general (baseline)<br>O: higher |
| High Prevalence Hypo-D Countries | P: much higher<br>O: much higher | P: much higher<br>O: much higher | P: higher<br>O: much higher | P: baseline<br>O: much higher |
| Low Prevalence Hypo-D Countries | P: lower<br>O: lower | P: lower<br>O: lower | P: higher<br>O: lower | P: baseline<br>O: lower |
| High Fish Diet Countries | P: lower<br>O: lower | P: lower<br>O: lower | P: higher<br>O: lower | P: baseline<br>O: lower |
| Dark-skinned Sub-populations | - | P: much higher<br>O: much higher | - | P: baseline<br>O: much higher |
| Tropic/Summer Lat < 40°N | Transmission | CFRs | Transmission | CFRs |
| General Summer Populations | P: lower<br>O: lower | P: lower<br>O: lower | P: lower<br>O: lower | P: baseline<br>O: lower |
| Quarantine, Distancing Measures | P: lower<br>O: lower | same as general (lower) | P: lower<br>O: lower | same as general (baseline) |
| High Prevalence Hypo-D Countries | P: higher<br>O: higher | P: higher<br>O: higher | P: baseline<br>O: higher | P: baseline<br>O: higher |
| Sun-Blocking Populations | - | P: higher<br>O: higher | - | P: baseline<br>O: higher |
| General Predictions | Transmission | CFRs | Transmission | CFRs |
| Elderly Sub-populations | - | P: much higher risk<br>O: much higher risk | - | P: higher risk<br>O: much higher risk |
| Pregnancy, Infancy Sub-populations | - | P: lower risk<br>O: lower risk | - | P: high risk<br>O: lower risk |
| D-reducing morbidities (A) | - | higher risk & coincidence | - | typical risk & coincidence |
| **D-impacted morbidities (B)** | - | higher risk and coincidence | - | typical risk & coincidence |
| **D-neutral morbidities (C)** | - | typical risk & coincidence | - | typical risk & coincidence |

#### Summary of Predictions versus Observations

**Model A:** 16 predictions match observed data; 3 predictions cannot be determined.

**Model B:** 14 predictions strongly contradict observed data; 2 may contradict data; 3 cannot be determined.

## Discussion

### Latitude Pattern and Outliers

During winter months in locations outside the tropics, the UV Index (UVI)^8^ reaches a maximum of 3 (Table 3) which is insufficient for the skin to produce Vitamin D [29].

**Table 3.** Maximum UV Index by month and latitude created from data published by the World Health Organisation [30].

| Country (City) | Latitude | winter |  |  | summer |  |  |  |  |  | winter |  |  |
| --- | --- | --- | --- | --- | --- | --- | --- | --- | --- | --- | --- | --- | --- |
|  |  | Jan | Feb | Mar | Apr | May | Jun | Jul | Aug | Sep | Oct | Nov | Dec |
| Russia (St Petersburg) | 60°N | 0 | 0 | 1 | 3 | 4 | 5 | 5 | 4 | 2 | 1 | 0 | 0 |
| USA (New York) | 41°N | 2 | 3 | 4 | 6 | 7 | 8 | 9 | 8 | 6 | 3 | 2 | 1 |
| Spain (Palma de Mallorca) | 39°N | 2 | 3 | 4 | 6 | 8 | 9 | 9 | 8 | 6 | 4 | 2 | 1 |
| Vietnam (Hanoi) | 21°N | 6 | 8 | 10 | 11 | 11 | 11 | 12 | 12 | 10 | 8 | 6 | 6 |
| Singapore (Singapore) | 1°N | 11 | 12 | 13 | 13 | 11 | 11 | 11 | 11 | 12 | 12 | 11 | 10 |
| Madagascar (Tananarive) | 19°S | 12 | 12 | 11 | 9 | 7 | 6 | 6 | 8 | 11 | 11 | 12 | 12 |
| Australia (Melbourne) | 37°S | 8 | 8 | 6 | 4 | 2 | 2 | 2 | 3 | 5 | 6 | 8 | 9 |
| New Zealand (Wellington) | 42°S | 7 | 7 | 5 | 3 | 1 | 1 | 1 | 2 | 4 | 6 | 7 | 8 |
| Falkland-Islands (Port Stanley) | 58°S | 5 | 4 | 2 | 1 | 0 | 0 | 0 | 1 | 2 | 3 | 5 | 5 |
|  |  | Jan | Feb | Mar | Apr | May | Jun | Jul | Aug | Sep | Oct | Nov | Dec |
|  |  | summer |  |  | winter |  |  |  |  |  | summer |  |  |

This broad pattern of UVI over season and latitude is well known to both correlate with and cause seasonal variations in serum levels of Vitamin D. The higher the latitude the higher the prevalence of vitamin D deficiency in winter months without supplementation. Outliers to this broad pattern require explanation.

The first severe COVID-19 outbreak in Europe was in Italy where vitamin D deficiency is one of the highest in Europe [31]; 76% (532/700) of women aged 60-80 were found to have 25(OH)D levels below 12 ng/ml [10] and hypovitaminosis D was 82% (14/17) prevalent in patients of long-term rehabilitation programmes[10,32].

In May the UK’s death toll overtook Italy’s. Belgium has the highest per capita deaths but a much smaller number of total deaths compared to Spain, Italy, UK and France [24]. Belgium was infected relatively late, on 11th March, with a relative “day zero” 49 days after reporting began so the severity of its outbreak is notable. A study to determine the vitamin D status of the population of Wallonia, Belgium (51.5°N) found that 45% (409/915) of the subjects were vitamin D deficient [33].

In Italy and Spain, infection and fatality rates dropped rapidly with the advent of spring and better weather. Spain had appeared to be on a worse trajectory than Italy, but the outbreak there occurred 3 weeks later than in Italy and appeared to have benefited from this delay and the effect of better weather earlier in its outbreak timeline.

The striking latitude relationship with severity revealed by the first analysis (Figure 3) is not explained by the timeline of the spread of SARS-CoV-2 infections (Figure 4). The first death occurred on 9th Jan 2020 in Wuhan, China. Systematic reporting began on 22 January once the severity of COVID-19 became apparent. Within two weeks, 35 neighbouring locations in Asia had reported at least one death or recovery, including Japan, Thailand, Hong Kong, and the Philippines, and one location in Australia (New South Wales). By the end of February, air travel had spread infections to 65 locations across Europe, North America, Oceania and South Asia. The majority of the 239 locations that ultimately reported infections had reported at least one recovery or death within the next three weeks.

The timeline of spread (Figure 4) was found to be broadly consistent with outbreak severity for locations above 30°N where locations infected early generally experienced more severe outbreaks. South of this line however, this was not the case. Multiple locations below 30°N reported infections early, some many weeks before severe outbreak in the north, and yet none progressed into severe outbreaks.

Available data for the four main northern outlier countries (Canada, Switzerland, Germany, Japan and South Korea) suggest that a common link is relatively low prevalence of vitamin D deficiency [11][34][35][36][37][38]. Canada and Switzerland were infected relatively late and therefore may have benefited from this delay. Canada in addition, may also benefit from a low population density. Canada is a very large country and a more detailed investigation of locations within it is merited. Inconsistencies in location definitions between deaths and recoveries data for Canada made it impossible for us to do a more detailed analysis. This raises sufficient concerns about the reliability of the data for Canada to justify excluding it until these can be resolved.

Japan was infected very early and has a very high population density but has the lowest incidence of vitamin D deficiency which is attributed to its high fish-content diet [39]. The reason behind the other countries is less clear but is presumed to be lifestyle, diet and a nationwide policy on supplementation. A lack of more recent data and issues with inconsistent testing methods and definitions make it difficult to assert this with certainty. Nonetheless, the broad pattern of serum 25(OH)D correlations with outbreak severity appears to hold consistently and accounts for outliers across the full latitude range.

Two locations worthy of comparison - Iran and Israel - lie in the relatively sunny latitude range 31 -32°N. Iran suffered a severe outbreak despite its lower latitude with an epidemic severity index of 3.4 and more than 2,500 deaths. In stark contrast, Israel reported just 12 deaths and 101 recoveries. Studies of vitamin D levels for the two countries reveal that Iran has a very high prevalence of vitamin D deficiency [40] whereas Israel is relatively low [41]. In Iran this is attributed to skin colour, religious full-body clothing and lifestyle.

Whilst there are far fewer studies of prevalence of vitamin D deficiency for locations in the tropics and southern hemisphere, the available data for identified outliers aligns with the predictions of the causal model:

Below 30°N, Egypt (26°N) appears as a potential outlier though it reported relatively late infection and its total numbers by 28th March are too small to draw conclusions. Further south, the Philippines (13°N) is a more distinct potential outlier though again total numbers are quite small but indicate higher fatality rates for hospitalised cases. The Philippines has noted serious concerns about vitamin D levels after a small study found that 58% (214/369) of randomly tested office workers in Manila were deficient [42]. Phillipine Dr Mark Alipio recently published a (preprint) retrospective multicentre study of 212 cases with laboratory-confirmed infection of SARS-CoV-2 from three hospitals in Southern Asian countries. Data pertaining to clinical features and serum 25(OH)D levels were extracted from the medical records. Alipio concludes, “vitamin D status is significantly associated with clinical outcomes (p<0.001). For each standard deviation increase in serum 25(OH)D, the odds of having a mild clinical outcome rather than a severe outcome were increased approximately 7.94 times; the odds of having a mild clinical outcome rather than a critical outcome were increased approximately 19.61 times,” indicating that, in COVID-19 patients, increased serum 25(OH)D level could improve clinical outcomes, and/or mitigate the worst (severe to critical) outcomes. Conversely, decreased serum 25(OH)D levels could worsen clinical outcomes.” [43].

Near the equator, Indonesia (0.8°S) has an ESI timeline similar to the Philippines but both trajectories are notably less severe than locations north of 30°N with a very low total number of fatalities (102). By the 11th April, Indonesia had reported 327 deaths, and 286 recoveries, indicating an approximate hospital fatality rate of 53% confirming its outlier status. Indonesia is an emerging economy and we were unable to find reliable data for vitamin D status in the general population, however a small study of 239 pregnant mothers in West Sumatra in 2019 reported first-trimester deficiency in 83% of cases. A retrospective study of 780 cases found 87.8% of Vitamin D insufficient COVID-19 cases and 98.9% of deficient cases ended fatally [44].

Brazil also reported its first deaths and recoveries outside the first seven weeks, but showed a steeper trend towards being an outlier in the southern hemisphere. By 11th April Brazil had reported 247 deaths and 173 recoveries indicating an approximate hospital fatality rate of 59% confirming it as an outlier in the south. Available data suggest that Brazil has a very high prevalence of vitamin D deficiency in elderly populations. Studies of two nursing homes in São Paulo found that residents had very low mean serum levels in winter which did not improve over summer. Mean ± standard deviation was reported as 37.6 (SD 29.9 nmol/L), compared to the recommended value >75.0 nmol/L [45]. It was noted that many of these individuals are from low income families that have little access to health services. Skin colour is likely to contribute significantly to vitamin D deficiency in Brazil, since it has a large population of black (8%) and mixed race (43%) ethnic groups [46].

### Causal Inference

The COVID-19 pandemic spread globally providing observational data with statistical power many orders of magnitude greater than a devised RCT that could be conducted even at national level. Striking patterns emerge directly from this statistical power that are so large they are evident without the need for sophisticated regression analyses. Global location data for 239 locations offers a vast data set that includes homogeneous and heterogeneous populations and subpopulations where latitude, weather conditions, skin colour, age, pregnancy and morbidity states are - in effect - randomly assigned by nature.

Our analysis considers data up to 9th April by which time there were 1.6M confirmed infections. Some estimates suggest this represents just 6% of actual infections suggesting our results could represent a globally distributed population of around 26M people, and this does not include those exposed to the virus which potentially raises the figure up to 1 BN.

Contrastive causal models allow us to distinguish a large set of predicted features from a multitude of predominantly *independent* exogenous variables driving them. This greatly increases statistical power^9^ through triangulation: the more independent root causes whose natural variation concurs with the model predictions, the narrower the possible range of explanations, and the less likely the model is to be wrong. This holds true even for hidden or unknown biases so long as the source of bias is different for each root variable and the tenet of randomisation does not hold true for some unknown reason.

Even if we discount possible confounders, age and underlying conditions, and exclude Canada, a large set of predictions remain that match those of the causal model and that do not match predictions made by the acausal “bystander” model.

Clear confirmation of the causal model is evident in the observed data as follows:

- Severe outbreaks with high fatality rates happened in general only in the northern hemisphere post-winter locations
- In general, outbreaks in tropical and southern post-summer locations were mild
- Northern outliers Canada, Germany, Japan and South Korea all correlate with known low prevalence of hypovitaminosis D relative to countries with severe outbreaks, presumed to be due to either high-fish containing diets or supplementation (actual cause is immaterial)
- Southern outliers: Philippines, Indonesia and Brazil with unusually high case fatality rates correspond with evidence of high prevalence of hypovitaminosis D in at-risk populations.
- Pregnancy and infancy are typically high-risk groups for most diseases yet in developed countries where these groups are routinely supplemented with vitamin D they have curiously not shown up as typically high-risk.
- Statistics for communities with genetically dark skin in the UK and US have confirmed case fatality rates twice the average rate.

The hypothetico-deductive method cannot verify a single model with absolute certainty, it can only falsify with absolute certainty. Einstein famously expressed this limitation when he said, “No amount of experimentation can ever prove me right; a single experiment can prove me wrong”.

Our proof considers two contrasting models, the verification of the causal model can be considered proof since the contrasting acausal model was falsified thus removing any remaining doubt about the result.

### Historical Evidence Suggests Causal Role of Vitamin D in Pandemic Prevention

Vitamin D was discovered five years after the 20th century’s deadliest pandemic, Spanish Influenza, caused by the avian virus H1N1, which infected one third of the world’s population (500M/1,8B) with a mortality rate of 10% to 20%. Vitamin D deficiency was rife: rickets was present in 80% to 90% of children living in northern Europe and north eastern US [7]. Lacking antibiotics, many died of secondary bacterial infections [47]. Sanatoria promoted fresh air and sunshine as a treatment that worked, but no-one knew why.

Between 1930 and 1950, Vitamin D fortification was commonplace and additives eradicated hypovitaminosis D. The practice was banned in 1950 on the grounds that uncontrolled dosing had led to instances of hypercalcaemia and hypovitaminosis D returned. The introduction of sunscreen creams containing UVB blockers in the 1970s further exacerbated this [7].

We note with interest that on the timeline of major influenza epidemics [48] there is a 37 year period from 1920 to 1957 where no new flu strains seem to appear and no new pandemics occurred (Figure 11). This coincides with the only known period during which the population at large was routinely supplemented with vitamin D. This historical observation in alignment with our analysis verifies the causal role of vitamin D supplementation in respiratory disease pandemic prevention.

**Figure 11.**
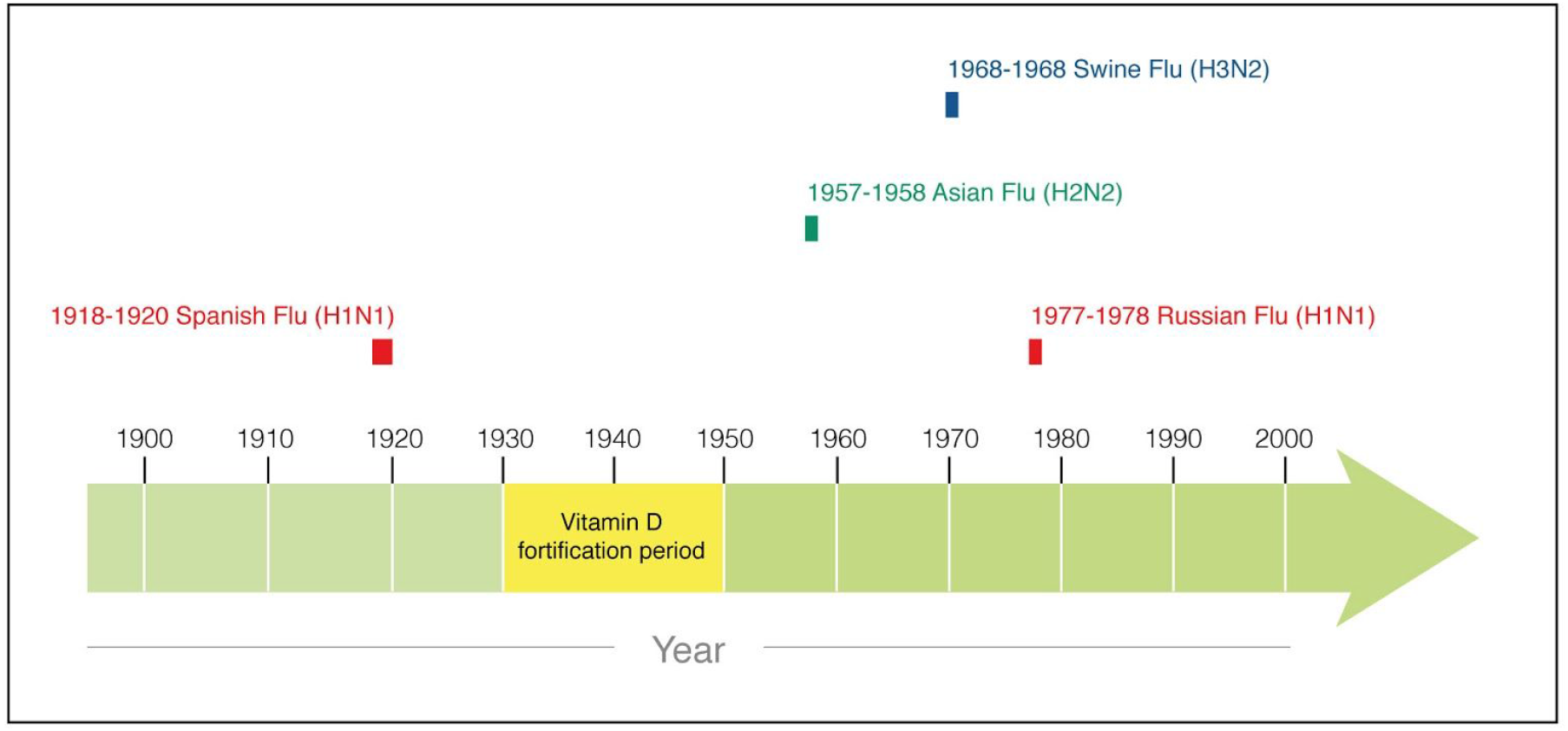
Influenza epidemic and pandemic strain outbreaks in the 20th century. No major outbreaks or major mutations occurred during vitamin D food fortification.

Recent concerns have been raised about the return and rise of rickets in the US and Europe

### Molecular Mechanisms Support Causal Inference

In coronavirus disease, a cellular receptor for the formerly identified SARS-CoV virus and the novel SARS-CoV-2 virus is the angiotensin-converting enzyme 2 (ACE2) [50][51].

ACE2 is a membrane-bound aminopeptidase which coregulates the renin-angiotensin system (RAS). The RAS plays a critical role in health and disease, including the regulation of the cardiovascular system and kidneys [52]. Here, angiotensin-converting enzyme (ACE) and ACE2 act in counterbalance to maintain finely tuned homeostatic processes such as blood pressure regulation and inflammation among others [53][54].

The respiratory tract is a major site of coronavirus infection and morbidity wherein SARS-CoV-2 causes COVID-19-related pneumonia and acute respiratory distress syndrome (ARDS)-like symptoms. Under normal conditions ACE2 is highly expressed in the lungs in order to balance the higher levels of Angiotensin II produced by ACE. Coronavirus studies have shown that viral replication can downregulate ACE2 [55]. A gradual depletion of ACE2 during the progression of SARS-CoV-2 infection can unbalance the RAS and lead to its activation. This eventually results in neutrophil infiltration, an exaggerated inflammatory host response, the cytokine storm, which is similar to ARDS and causes injury to the lungs.

ACE2 has been shown to be protective against ARDS in animal models where higher ACE2 levels reduced the severity of ARDS [56]. Understandably, ARDS can be prevented by treatment with an angiotensin II receptor antagonist RAS inhibitor, losartan, which has been shown to upregulate ACE2 [57].

RAS activation has proinflammatory and profibrotic effects [58] that are present in many chronic diseases including cardiovascular disorders, diabetes, and lung fibrosis. RAS activation in lung fibrosis can be induced by chronic vitamin D deficiency [59]. In addition, ARDS is also strongly associated with vitamin D deficiency [60], which occurs in 90% of cases and correlates with poor disease outcome [61]. These observations imply that vitamin D sufficiency can prevent RAS activation and thus ARDS.

Vitamin D (calcitriol) acts by negatively regulating the RAS, modulating expression of its component members: renin, ACE and ACE2. 1,25-dihydroxyvitamin D3 suppresses renin gene transcription by blocking the activity of the cyclic AMP response element in the renin gene promoter and the ACE/Ang II/AT1R cascade [62][63] as well as inducing ACE2/Ang-(1 -7) axis activity [64].

In summary, vitamin D inhibits RAS activity by decreasing the renin input to the RAS upstream, while increasing ACE2 downstream and thus protects the critical balance of the system.

In addition to the mechanisms outlined above, vitamin D sufficiency protects against infection via binding to the vitamin D response element in many gene promoter regions resulting in gene expression changes. This leads, for example, to a decrease in proinflammatory cytokines and an increase in antiviral and antibacterial peptides such as defensin *β 2* and cathelicidin [65]. HIV studies have also revealed that it promotes an anti-inflammatory response by inhibiting the maturation of dendritic cells (DCs), downregulating antigen presenting molecules (MHC-class II), costimulatory molecules (e.g., CD40, CD80, and CD86), and pro-inflammatory cytokines (e.g., IL-12 and IL-23) in addition to enhancing anti-inflammatory cytokine (IL-10) and T-cell inhibitory molecule (PD-1) [66].

The known molecular mechanisms of calcitriol are in alignment with our causal analysis as well as the historical evidence, and corroborates a beneficial causal role of vitamin D in COVID-19 outcomes.

### Emerging Clinical Research Evidence

Since our early findings and call to action on 12th and 19th March, corroborating observational studies have appeared in preprint. Several show a significant correlation between hypovitaminosis D in study subjects and the severity of their COVID-19 outcome:

- a retrospective multicentre Philippine study of 212 cases concludes that vitamin D status is significantly worse in severe COVID-19 patients [43].
- another retrospective study from India with 176 cases of elderly (>=60 years) found that 86% of severe COVID-19 cases were deficient of vitamin D (25(OH)D<30ng7ml) [67].
- a smaller study from USA shows that vitamin D insufficiency (serum 250HD < 30ng/ml) is 84.6% prevalent in ICU admitted COVID-19 patients [68]
- from Indonesia a larger retrospective cohort study of 780 cases found 87.8% of Vitamin D insufficient COVID-19 cases and 98.9% of deficient cases ended fatally [44].
- a study in European countries appeared to establish a correlation that the lower the mean vitamin D level (25(OH)D), the higher the number of COVID-19 cases/million [69].

These observational studies show significant correlations between hypovitaminosis D and COVID-19 severity in complete alignment with our results, however none serves as proof of causation.

One study we should discuss claims no link between vitamin D concentrations and risk of COVID-19 infection either overall or between ethnic groups [70]. The study correlates a small sample of recent COVID-19 infection test results with serum 25(OH)D concentration data sampled between 2006 and 2010, which assumes serum levels are stable both seasonally and over 10-14 years. Additionally, evidence has pointed to vitamin D deficiency impacting morbidity and mortality, not infection risk, as this study suggests. Further discussion is not merited because of these methodological concerns.

As an independent source of evidence, we note with interest that severe vitamin D deficiency is prevalent in Kawasaki disease [71], a rare systemic inflammation of the blood vessels with unknown origin. This appears to occur in younger COVID-19 patients with 30-fold increase in incidence [72]. This is consistent with the predictions of the causal model.

Vitamin D supplementation strategy for future COVID-19 patients may vary substantially based on underlying clinical conditions and needs to be medically evaluated. Moreover, we encourage the medical and healthcare communities to clarify for their subspecialties whether solely nutritional doses and approaches of vitamin D are adequate for maintaining vitamin D sufficiency, according to individual needs, as some nutritionists have assumed and advised in the media.

Future research is needed to understand whether vitamin D is able to reverse fibrotic pathological changes observed in COVID-19 survivors, since multiple studies suggest a beneficial effect of vitamin D in reversing fibroses [73-77].

Morbidity can be influenced by many factors, all sources of noise, so the fact that our analysis yielded such unambiguous results implies that vitamin D status is likely the dominant contributor to COVID-19 outcomes.

We encourage investigations into the roles of vitamin C, zinc, and vitamin B12 in a COVID-19 context as these are widely recognised also to contribute to a healthy host immune response, as well as Vitamin K which is involved in blood coagulation pathways, which appear to be key aspects in COVID-19.

### RCTs versus Observational Methods

RCTs are widely considered the ‘gold standard’ for clinical trials but there is “significant concern that the lay public and sometimes researchers put too much trust in RCTs over other methods” and that “their status is exaggerated” [78,79]. Disturbingly, findings from flawed RCTs have overturned policy decisions when they contradicted better evidence from observational study findings [80].

There is a widespread misunderstanding that RCTs prove cause, but RCTs only *permit* the inference of cause from data *if certain criteria are met* and unfortunately they rarely are. A study of ten of the most cited RCTs worldwide found none met minimum standards and called into question their results [81]. RCTs have many major drawbacks: multimillion dollar costs, length of time to design and run, profound ethical issues, and strong industry bias. On top many other methodological flaws, trial participants rarely represent the target treatment population and findings often fail to be reproduced in wider clinical practice. A 2015 study found that patient samples in the majority of RCTs in cardiology, mental health, and oncology were not representative of patients in real world settings [82]. RCTs continue to be demanded as “proof of cause” despite representing relatively weak evidence.

In studies of all types, population size significantly determines the statistical power, strength and credibility of evidence. RCTs are fundamentally size-limited by cost and practicality. Observational methods have no such restriction and as they develop in sophistication, and as data become more available to apply them in appropriate contexts, they justify higher rank in the hierarchy of evidence. The hypothetico-deductive method used in this paper is rarely employed in medicine, yet is the basis of centuries of advancements in the field of physics. We hope that our example here will encourage its adoption more widely in future medical research.

For further reading see *Appendix 2 - RCTs, Causal Inference and the Hierarchy of Evidence*.

## Conclusions

Our novel causal inference analysis provides high-quality evidence supporting the hypothesis that vitamin D plays a causal role in COVID-19 outcomes via modification of host responses to SARS-CoV-2.

The size and scope of available data used in our analysis is unprecedented with almost 240 global reporting locations with 1.6 million confirmed infections and covering a population of circa 1 billion, making it potentially one of the highest power causal inference studies ever performed. Using a hypothetico-deductive method, 100% of testable predictions verified the causal model and falsified the acausal model, providing compelling evidence in favour of a causal relationship in advance of RCTs. The application of observational causal inference methods is relatively new in medicine, but their many advantages in time, cost, study size and strength of evidence suggests they should rank higher in the hierarchy of evidence and play a major role in the future of medical research.

Since vitamin D is implicated in the pathogenesis of numerous diseases, humankind could significantly benefit from a global effort to collect reliable, high quality, standardised data for 25(OH)D levels for future CI applications.

In addition, historical evidence is highly consistent with a causal protective role for vitamin D in respiratory virus infections. There is a complete absence of new major epidemics and pandemics caused by respiratory viruses in the twentieth century during the period when vitamin D supplementation was applied at population level. This strongly implies that vitamin D has played - and indeed does play - an important causal role in seasonal respiratory disease susceptibility and outcomes.

The RAS dependent and independent molecular evidence presents a feasible explanation for the underlying mechanisms that explain the beneficial preventive role of vitamin D in COVID-19 outcome and demonstrates how a vitamin D modified host response can prevent pathological changes. Finally, we would note that emerging preliminary clinical research data show a significant correlation between hypovitaminosis D and severity of COVID-19 outcomes, which is in complete alignment with our conclusions.

Based on the evidence from our causal inference analysis, we strongly recommend vitamin D supplementation to ensure sufficiency across the global population. Vitamin D prophylaxis as a host response modifier, offers a widely available, low-risk, highly-scalable, and cost-effective strategy to prevent local outbreaks and the second wave anticipated later this year, and end the COVID-19 pandemic. Vitamin D prophylaxis can be implemented immediately in advance of vaccines and medications under development, and moreover, offers long-term added value for the general health status of the population.

The timely implementation of vitamin D supplementation programmes worldwide is critical, and initial priority should be given to those who are at the highest risk, including the elderly, immobile, homebound, BAME and healthcare professionals. Vitamin D supplementation has to follow nationally accepted recommendations under the advice of qualified health professionals, to reach serum 25(OH)D level sufficiency whilst avoiding risk of overdose and hypercalcaemia.

Ensuring population-wide vitamin D sufficiency could mitigate seasonal respiratory epidemics, decrease our dependence on pharmaceutical solutions, reduce hospitalisations, and thus greatly lower healthcare costs while significantly increasing quality of life.

## Data Availability

Data and code are available online at github.

https://github.com/gruffdavies/GD-COVID-19

## Footnotes

## Acknowledgements

We would like to thank Professor Andrea Giustina and Dr Thomas Hiemstra for advice and feedback during the process of writing this paper; Gordon Shotwell for early feedback on the first and second preprint submission drafts; Eoin Norton for generous graphic design assistance. Dr Davies would like to thank Andrew Lewis for first introducing him to the surprising range of beneficial actions of Vitamin D in 2019, Susan Greenwood for kind support, Heather Briant and Simon Potter for patience and understanding during a testing time. Dr Garami would like to thank Tibor Olajos for calling to his attention at the beginning of March the novel fact that ACE2 serves as a receptor for SARS-CoV-2. Dr Byers would like to thank Joshua Lau for his advice on the pharmacological management of vitamin D deficiency in adults.

## About the authors

**Dr Gareth Davies** is a British physicist and tech entrepreneur (BSc, Physics; PhD, Medical Physics, Imperial College). Although he is not currently affiliated with Imperial College as a research scientist, he has returned numerous times in recent years to give guest talks on AI and Entrepreneurship. He has more than three decades of experience of complex data analysis, systems modelling, software engineering and machine learning. He is a recognised pioneer in several fields, and in September 2019 he was named as one of World’s Top 50 Innovators by Codex at the Royal Society in London.

**Dr Attila R Garami** is a Hungarian medical doctor (MD, Albert Szent-Györgyi Medical University) and holds a PhD from the Doctoral School of Multidisciplinary Medical Sciences, University of Szeged, Hungary. He has a wide range of expertise in tropical diseases, immunology, biochemistry and molecular biology from the Max Planck Institutes in Tübingen, Germany, where he earned a reputation as an outstanding research fellow. Later, as an EMBO Long-Term Fellow, he specialized in growth- and cancer-related signaling from Friedrich Miescher Institute for Biomedical Research, Basel in Switzerland. Afterwards, he participated in the pioneering establishment of the multidisciplinary biomarker field in the pharma environment. He lives in Switzerland and has been working as a senior consultant in the field of innovative biomarkers for 15 years.

**Dr Joanna Byers** is a British medical doctor (MBChB, University of Birmingham) with 5 years of clinical experience across primary and secondary care sectors. She holds a diploma in Global and Remote Health Care; Leadership and Innovation from the University of Plymouth. Her current focus and interests are the design and implementation of effective preventative health services using a person-centred approach to care, and she is currently undertaking an MSc in Occupational Therapy with the University of Essex. She also facilitates Public Health teaching at the University of Cambridge Medical School.

## Background to this paper

In response to the urgency of the unfolding crisis the authors separately began to research and publish vitamin D hypotheses. Dr Garami wrote a BMJ Rapid Response letter published on 12th March [3]. We wrote to him and were delighted when he agreed to our invitation to collaborate. We published early research findings on 19th March [5] outlining the detailed evidence supporting a beneficial role for vitamin D in the prevention and treatment of COVID-19, and calling for hospitals to test serum levels, treat deficiency and report data and outcomes. Response to the report was highly polarised. Those critical of it were quick to dismiss citing “correlation isn’t causation”. The report contained direct and indirect evidence for cause but lacked a formal framework justifying causal inference which has slowed the adoption of this critical information. A causal inference framework has a great deal of value to add beyond the current pandemic. Vitamin D has received a great deal of renewed attention in recent years, but a beneficial role for it in disease states has been controversial [82]. Vitamin D deficiency in association with many serious diseases has raised the question of whether it is a bystander, simply a general marker of illheath, or plays an active role in the cause and progression of diseases. The lack of standardised serum level testing and a long history of research and clinical trials with significant design flaws adds significant noise and confusion to the body of research making it difficult to find clear answers even when present. We hope that the causal model framework presented in this paper will help to resolve some of these controversies.

## Appendix 1 - Background on Causal Inference

A new science of “causal inference” (CI) emerged in recent years from the field of artificial intelligence and was introduced to health research around the turn of the millenium [83]. CI is an extension to statistics that defines causation mathematically and allows it to be determined if causation can be inferred from observational data. If so, the size of the causal effect can be calculated.

CI employs causal diagrams (models) which are mathematically and graphically represented by “directed acyclic graphs” (DAGs). These network-like diagrams of connected nodes declare the flow of causes-to-effects in the model, and CI’s new symbolic language makes it possible to mathematically express and answer the causal questions we wish to pose. Formal rules dictate how DAGs are created, manipulated and interpreted soundly, and CI gives ways to test models for correctness and identify potential flaws. CI formally distinguishes *seeing* from *doing*, (adding a “do-operator”) which allows for a new, highly-succinct definition of “confounding” as: the difference between *seeing X* and *doing X* on an outcome (Y). If *seeing* and *doing are* equal in outcomes, there is no confounding, otherwise there is. Prior to CI, statistics has been unsuccessful in clearly, succinctly and correctly defining confounding. The nature of confounding was ironically itself confounding.

**Figure 12.**
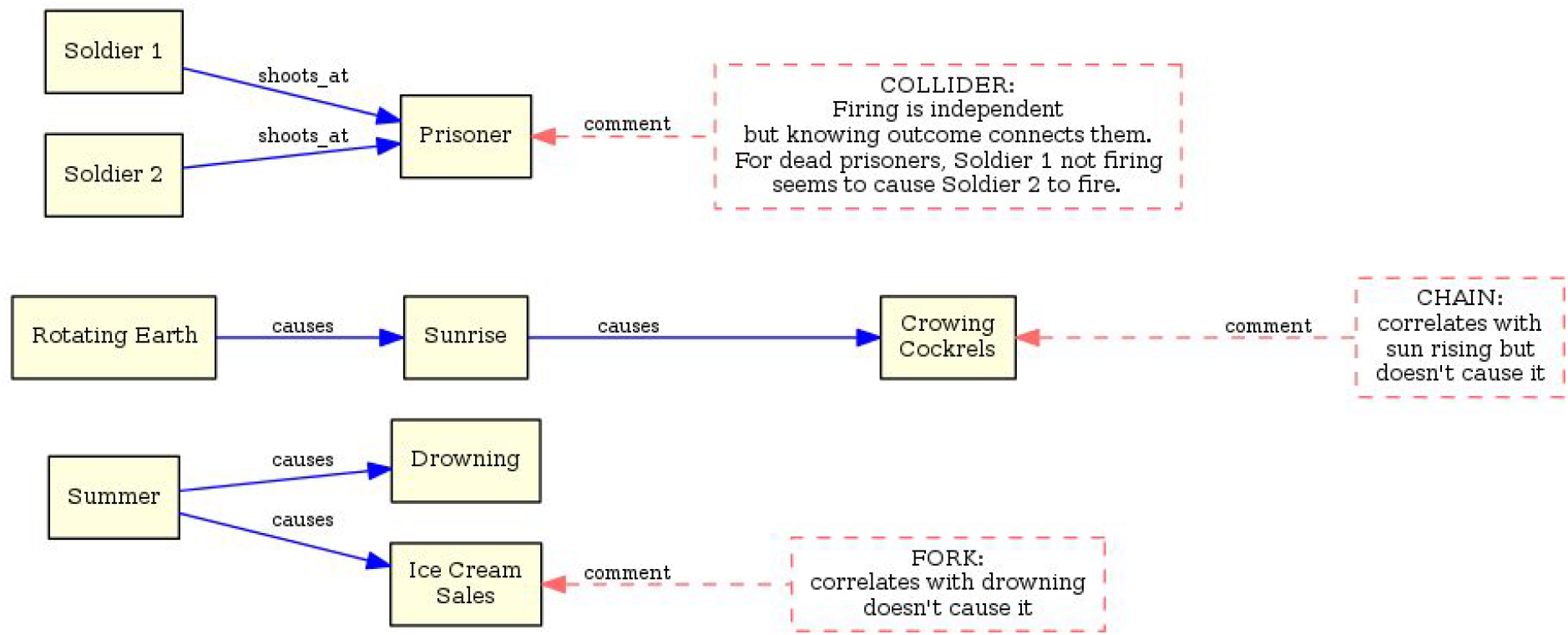
examples of sources of confounding for the three types of causal relationships in DAGs: “collider (top), “chain” (middle) and “fork” (bottom).survi

The purpose of a causal model is to *explain* the underlying process generating the observed data. The process of drawing and iterating DAGs brings clarity and interpretability to what can otherwise be highly-confusing data. Most importantly, models generate predictions which are testable against observed data and can be verified or falsified. Model relationships can be expressed mathematically to calculate the expected size of a causal effect purely from observed data with remarkable accuracy [84]. Models help to identify spurious correlations and also show when inference is not possible. A credible causal model must first be defined then analysed to identify potential confounding variables, and any variables that may appear to be confounders but are not. Confounding variables can be “controlled for”^10^ enabling the deconfounding of statistics. Models help to identify variables that should not be controlled for (colliders), since doing so may inadvertently introduce selection bias. CI gives us the power to interpret the size of causal effects from correlations in observational data.

*The Book of Why* (Judea Pearl) is an excellent, accessible book on Causal Inference intended for lay readers.

Further reading (technical):

- *Causal Inference In Statistics - A Primer*(Judea Pearl, Madelyn Glymour, Nicholas P. Jewell)
- *Causality* (Judea Pearl)
- *Causal Inference for Statistics, Social, and Biomedical Sciences: An Introduction* (Guido W. Imbens, Donald B. Rubin)

## Appendix 2 - RCTs, Causal Inference and the Hierarchy of Evidence

Since James Lind conducted the first clinical trial proving that citrus fruit cured scurvy in 1753, RCTs have enjoyed many outstanding successes, and those of the late 20th century especially have led to its adoption as the “gold standard” for determining cause and estimating Average Treatment Effect (ATE) in medicine.

RCTs are merely one of many causal inference methods science has developed. Methods using observational data are increasingly used in medicine, but have been slow to be accepted. The preferential status of RCTs above other methods is being questioned - and rightly so. Modern RCTs are the subject of much deserved criticism and debate, not least because of their exorbitant costs^11^, the time they require to design and run, profound ethical issues, and that many are now funded and run by pharmaceutical companies with major conflicts of interest. In 2014, the UK parliament’s Public Accounts Committee issued a damning report [86] finding that “information is routinely withheld from doctors and researchers about the methods and results of clinical trials on treatments currently prescribed in the United Kingdom”. Publication bias is not limited to privately funded RCTs. Negative findings are less likely to be reported or accepted by journals even if submitted [87] making it impossible to objectively judge overall results. A 2017 systematic review of RCTs raised concerns about lack of transparency and comprehensive data [88].

Like all causal inference methods, RCTs *permit* the inference of cause from data *if certain criteria are met*. Unfortunately, in the case of RCTs, they rarely are. A comprehensive analysis of ten of the most influential and cited RCTs worldwide found that they did not meet minimal quality standards - with multiple sources of unexamined and undeclared sources of bias - and called into question the robustness of their results [81].

There is significant concern that the lay public and sometimes researchers put too much trust in RCTs over other methods and that their status is exaggerated [78,79]. This especially concerning when findings from flawed RCTs have preferentially influenced policy over those from correct observational study findings [80].

There seems to be a poor appreciation of the frailties and limitations of RCTs. Blinding matters for unbiasedness but is often missing or compromised. Belief that “randomisation” magically removes bias, is the only way to do so, and that RCTs are proof of cause - none of which is true - has a disturbingly superstitious quality to it. Randomisation does *not* balance confounders in any single trial, and in fact, the smaller the trial the higher the chance of it doing the exact opposite. Regression techniques are widely used to adjust experimental data presuming this improves data quality, but without checking fundamental prerequisites are met, and since randomisation does not justify the model, almost anything can happen [89].

Unsurprisingly, the findings of many modern RCTs fail to be reproduced when applied in wider clinical practice. On top many other methodological flaws contributing to this, trial participants rarely represent the population intended for treatment. A 2015 review RCTs found that patient samples in the majority RCTs in cardiology, mental health, and oncology were not representative of patients in real world settings [90]. Yet, RCTs continue to be demanded as “proof of cause” when they are patently no such thing.

The myopic attachment to RCTs in medicine and its distrust of observational methods is as strange as it is surprising. Many observational methods have an outstanding history of success in other fields. The hypothetico-deductive method employed in this paper has been the mainstay of physics for centuries. It has been used to confirm Einstein’s theory of relativity, Newton used it to prove the moon causes the tides, it helped determine the structure of the atom, develop quantum theory, discover the Higgs boson, and has been the source of some of the biggest paradigm shifts in mankind’s history overthrowing millennia of religious belief. Copernicus used its power to move the very Earth (figuratively) and Darwin, with his theory of Evolution, used it to cut God from story of creation.

RCTs capably answered many simple medical questions of the 20th century, but few modern RCTS are conducted with sufficient care to represent consistent, reliable evidence of causation. This is not to suggest that RCTs have no value or place in science. It is the reification of RCTs that is the problem, not RCTs themselves. They are a victim of their 20th century success. In a manner reminiscent of the Peter Principle from management, RCTs have been promoted to their level of incompetence. The solution is not to dispense with them, but to demote them back to a more appropriate level. The idea that RCTs deserve rank above observational methods is a religious belief - not an evidence-based one. So long as magical thinking surrounding RCTs continues to insist that they be the only arbiter of cause, the vast cost to society will not merely be economic but measured in human lives.

Observational methods offer the potential to discover new causal relations, and confirm or falsify those suggested by RCTs.

Change though slow, has begun: policy decisions concerning specific influenza vaccines based on RCT efficacy estimates have been overturned in recent years thanks to observational studies showing they did not in fact work [91].

## Footnotes

1 JH CSSE COVID-19 data for deaths in Canada is reported for 15 locations but recoveries in Canada are reported for the whole country, requiring that we aggregate the deaths into a single figure.

2 Readers with a non-mathematical/physics background: https://en.wikipedia.org/wiki/Inverse_problem

3 Red category membership lines are also labelled “is” so these can be printed in black and white.

4 Linearised form of ESI raises ten to the absolute ESI value so that geobubble areas are directly comparable with the number of deaths.

5 ESI is useful for quantifying and quickly determining severity and outliers in a single statistic but as with any single statistic, the devil is in the detail beneath.

6 Any challenges to the assertion of independence can be handled by modifying the diagram appropriately or similarly considering the expected nature of the impacts.

7 Sophisticated techniques using DCGs have been developed recently that can handle cyclic dependencies.

8 The UV Index is a convenient standard for expressing UV radiation levels.

9 Although this is a qualitative analysis answering a binary question, the concept of statistical power is still important even though we do not calculate a numerical value for it.

10 To “control for a variable” is to hold it constant, or within a range, in order to remove it as a possible cause for some outcome further down the causal chain. This can clarify if another variable is truly at cause. Care must be exercised choosing control variables: controlling inappropriate variables may generate statistical bias incorrectly implying cause where there is none.

11 A study of 138 trials of 59 new therapeutic agents approved by the FDA from 2015 to 2016 revealed costs ranged from less than $5M to $347M with the median estimated trial cost of $19M [85]. One $5M trial had no control group and studied fewer than 15 patients.

